# Atrophy in multiple system atrophy relates to mitochondrial and oligodendrocytic processes

**DOI:** 10.1101/2025.01.22.25320961

**Authors:** Lydia Chougar, Christina Tremblay, Aline Delva, Marie Filiatrault, Andrew Vo, Justine Y. Hansen, Asa Farahani, Bratislav Misic, Parsa Khalafi, Charles-Etienne Castonguay, Guy Rouleau, Jean-Christophe Corvol, Marie Vidailhet, Bertrand Degos, David Grabli, Stéphane Lehéricy, Alain Dagher, Shady Rahayel

**Affiliations:** The Neuro - Montreal Neurological Institute and Hospital, McGill University, Montreal H3A 2B4, Canada; Sorbonne Université, Institut du Cerveau - Paris Brain Institute - ICM, AP-HP, CNRS, Inserm, Hôpital de la Pitié Salpêtrière, DMU DIAMENT, Department of Neuroradiology, F-75013, Paris, France, Paris, France; Center for Advanced Research in Sleep Medicine, CIUSSS-NÎM – Hôpital du Sacré-Coeur de Montréal, Montreal H4J 1C5, Canada; Department of Neuroscience, University of Montreal, Montreal H3T 1A4, Canada; Sorbonne Université, Institut du Cerveau - Paris Brain Institute - ICM, AP-HP, CNRS, Inserm, Hôpital de la Pitié Salpêtrière, Department of Neurology, F-75013, Paris, France, Paris, France; Dynamics and Pathophysiology of Neuronal Networks Team, Center for Interdisciplinary Research in Biology, Collège de France, CNRS, UMR7241/INSERM U1050, Université PSL, Paris, France; Department of Neurology, Avicenne University Hospital, Sorbonne Paris Nord University, NS-PARK/FCRIN network, Bobigny, France; Department of Medicine, University of Montreal, Montreal H3T 1A4, Canada

## Abstract

**Objective:** To investigate the gene expression and neurobiological underpinnings of brain atrophy in multiple system atrophy (MSA) using imaging transcriptomics and PET-based molecular annotation.

**Methods:** We derived brain atrophy measurements from the T1-weighted MRI scans of 65 patients with MSA and 181 age- and sex-matched healthy controls. Using postmortem data from the Allen Human Brain Atlas, partial least square (PLS) regression was used to identify gene expression components associated with atrophy. Gene enrichment analyses were performed to investigate the biological processes with enriched genes in regions showing atrophy. Annotation mapping was used to identify the neurochemical systems whose density maps matched atrophy in MSA.

**Results:** Atrophy in MSA was found to primarily affect deep brain regions, including the cerebellar white matter, pons, putamen, olive nuclei, and substantia nigra. PLS regression on deep brain region atrophy identified two gene expression latent variables, explaining 27.5% of the covariance. Regions with greater atrophy overexpressed genes related to the mitochondrial respiratory chain, particularly proton transmembrane transport and complex I assembly. In addition, cell type analysis revealed that regions with atrophy overexpressed genes related to oligodendrocytes. These patterns were distinct from those found in Parkinson’s disease. Atrophic regions in MSA also displayed less serotonin and GABA receptors and more acetylcholine and noradrenaline receptors.

**Interpretation:** Regions showing atrophy in MSA show specific features, namely overexpression of genes related to mitochondrial function and oligodendrocytes and distinct neurochemical patterns. This study highlights some of the biological and neurochemical mechanisms underlying selective vulnerability of brain regions in MSA.

## Introduction

Multiple System Atrophy (MSA) is a rare, rapidly progressing neurodegenerative disease with a poor prognosis, characterized by autonomic failure and varying degrees of motor impairment, including poorly levodopa-responsive parkinsonism in the parkinsonian variant (MSAp) and predominant cerebellar symptoms in the cerebellar variant (MSAc).^1^ MSA is classified as a primary synucleinopathy, along with Parkinson’s disease (PD) and Dementia with Lewy bodies, and is characterized by the accumulation of misfolded α-synuclein.^2,3^ In MSA, α-synuclein aggregates mainly within the cytoplasm of oligodendrocytes, forming glial cytoplasmic inclusions, which preferentially accumulate in the substantia nigra (SN) and striatum in MSAp, and in the cerebellum, pons, and olive in MSAc. Striatonigral degeneration and olivopontocerebellar atrophy often overlap in MSA as the disease progresses.^2,3^ Although considered a sporadic disease, rare familial occurrences have been reported, with variants at the *SNCA*, *MAPT,* and *COQ2* loci associated with the disease.^4,5^

Neuroimaging techniques such as magnetic resonance imaging (MRI) and positron emission tomography (PET) provide *in vivo* biomarkers sensitive to brain alterations.^6,7^ However, these imaging-derived phenotypes remain indirect proxies of the biological processes underpinning disease pathophysiology. Imaging transcriptomics is a novel method investigating the relationships between brain anatomy and gene transcriptional activity.^8^ In PD, brain iron deposition has been shown to relate to genes involved in heavy metal detoxification, synaptic function, and nervous system development and to map onto the brain distribution of astrocytes and glutamatergic neurons.^9^ Imaging transcriptomics also revealed that the progression of cortical atrophy over two and four years in PD was more prominent in regions overexpressing genes related to mitochondrial^10^ and synaptic functioning^11^ and less prominent in regions containing more oligodendrocytes and endothelial cells.^11^ In isolated rapid eye movement sleep behaviour disorder (iRBD), a prodromal stage of synucleinopathies, genes involved in mitochondrial function and macroautophagy were most strongly expressed in regions showing cortical thinning.^12^ To further characterize the biological underpinnings of neurodegenerative diseases and overcome limitations imposed by the collection of large datasets of PET-imaging tracers, spatial mapping annotation approaches have allowed the investigation of the spatial resemblance between brain patterns and specific density maps from individual receptors.^13^ In iRBD, brain atrophy has been shown to overlap with dopamine, serotonin, and noradrenaline density maps.^12^ To date, these approaches have never been used to characterize the biological underpinnings of brain damage in MSA. Such results could be helpful in identifying novel therapeutic targets in MSA.

Our objective was to quantify the pattern and extent of brain atrophy in MSA patients and its subtypes and apply imaging transcriptomics and PET-based spatial mapping annotation approaches to determine the biological characteristics underlying this atrophy. First, we characterized the pattern of brain atrophy in MSA patients. Next, we used multivariate analyses to derive gene expression components associated with the atrophy, using postmortem data from the Allen Human Brain Atlas. We applied gene enrichment analyses to uncover the biological processes enriched in the regions showing atrophy in MSA. We further used an annotation mapping approach to identify the neurotransmitters, receptors, and binding sites whose density maps spatially matched the atrophy pattern in MSA. To ensure that the resulting patterns were specific to MSA, we repeated the same analyses in a group of patients with PD. We hypothesized that brain atrophy in MSA targets regions with specific gene expression and neurochemical characteristics similar to other synucleinopathies in terms of mitochondrial dysfunction but distinct in terms of cell type involvement.

## Methods

### Participants

This is a retrospective single-center study of 69 patients with clinically established MSA enrolled prospectively: 1) between 2007 and 2012 at the Paris Brain Institute (ICM) as part of two research protocols (Genepark (LSHB-CT-2006-037544) and BBBIPPS (DGS 2006/0524)) and 2) between 2013 and 2020 in the movement disorders clinic at the Pitié-Salpêtrière University Hospital, Paris, as part of their diagnostic work-up. The diagnosis of clinically established MSA was retrospectively confirmed by a movement disorders neurologist, based on international diagnostic criteria.^1^ Participants were excluded if they had other neurological disorders, including stroke or brain tumor on MRI or if MRI findings were inconsistent with the clinical diagnosis (e.g., midbrain atrophy suggestive of progressive supranuclear palsy seen in a patient with a clinical diagnosis of MSA). Based on neurological examination, patients were clinically classified as the parkinsonian subtype (MSAp) when the parkinsonian syndrome predominanted, cerebellar subtype (MSAc) when the cerebellar syndrome predominanted, or mixed subtype when both syndromes were equally present.

From the same sites, we recruited 110 age- and sex-matched healthy controls without a history of neurological or psychiatric disorders as well as 80 controls enrolled as part of the Parkinson’s Progression Markers Initiative Database (PPMI).^14^ Data used in this work were obtained in February 2024 from the PPMI data, RRID:SCR 006431.

The local institutional review board approved the study (Genepark: CPP Paris II, 2007-A00208-45; BBBIPPS: CPP Paris VI, P040410–65-06; Parkatypique:CPP Ile-de-France VI08012015). Participants gave written informed consent.

### MRI acquisition

Participants were scanned using different 3T MRI scanners, namely a Siemens TRIO (Siemens Healthineers, Erlangen), Siemens SKYRA, General Electric (GE) SIGNA (GE Healthcare, Chicago, IL), and a 1.5T GE OPTIMA scanner. The acquisition included a three-dimensional gradient-recalled echo T1-weighted sequence (Tables S1 and S2 and PPMI imaging protocols).

### MRI processing

Figure 1 provides an overview of the pipeline. FreeSurfer (v7.1.1) was used for cortical parcellation and volumetric segmentation of the T1-weighted images.^15^ Maps passing quality control were segmented to derive 68 bilateral cortical thickness measurements from the Desikan-Killiany atlas and 14 volume measurements from the bilateral subcortical structures (putamen, caudate, pallidum, thalamus, nucleus accumbens, amygdala, hippocampus). The brainstem was segmented into its subregions to extract volumes for the midbrain, pons, medulla, and superior cerebellar peduncles using the brainstem toolbox within FreeSurfer.^15^ In addition, the cerebellum was segmented into 28 volumes, including lobules and cerebellar white matter, using CerebNet (v.2.1.2) ^16^ and the Schmahmann atlas.^17^ Volume measurements were normalized by the total intracranial volume derived from FreeSurfer to correct for head size.

**FIGURE 1.**
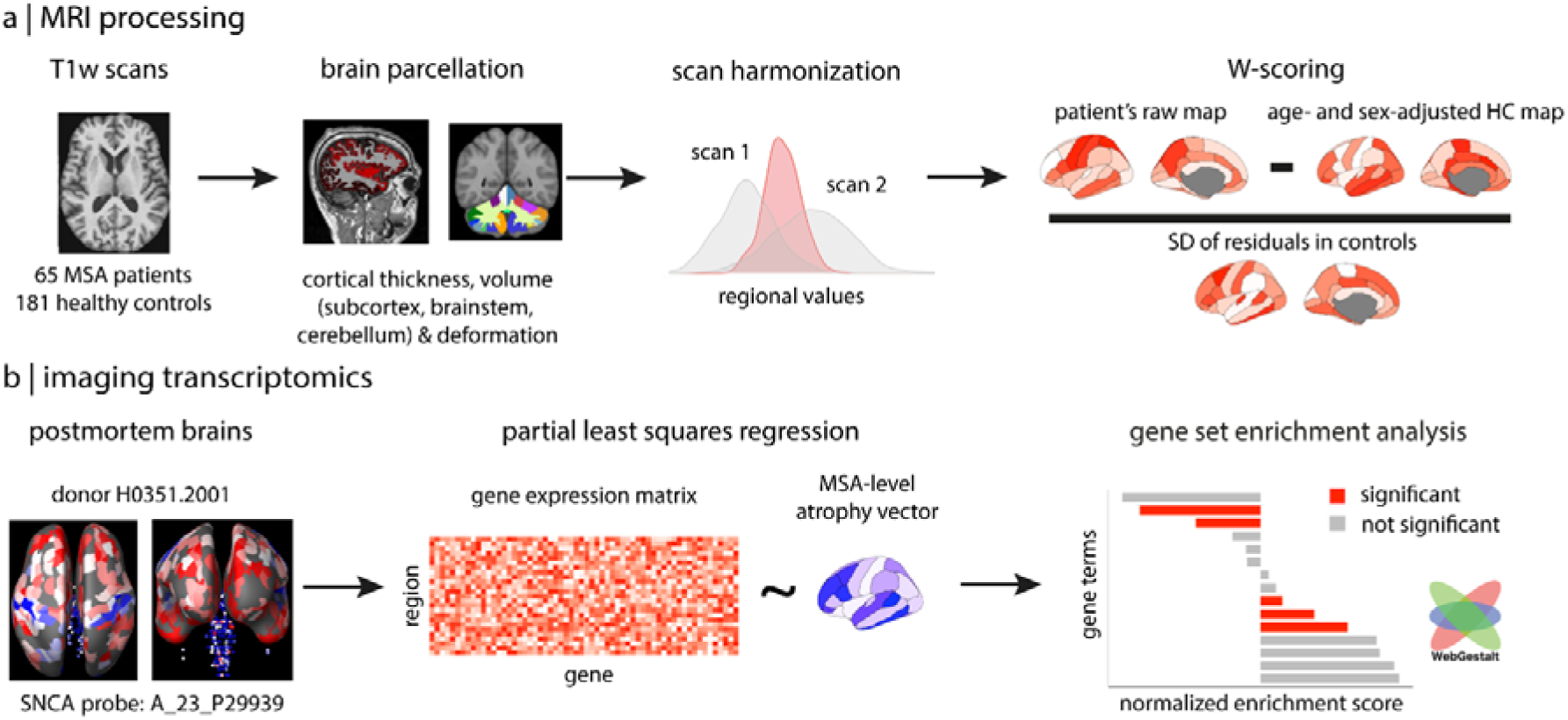
Workflow for MRI processing and imaging transcriptomics analysis. (a) MRI processing was done using T1-weighted scans from MSA patients and healthy controls. Processing maps were parcellated to compute cortical thickness, volume, and deformation metrics. Scan harmonization was applied to correct for inter-scanner variability. Regional values were converted to W-scores by subtracting age- and sex-adjusted healthy control values and dividing by the SD of residuals in controls. (b) The imaging transcriptomics analyses involved extracting regional gene extraction profiles from postmortem brain data. Partial least squares regression was used to associate the MSA-level brain atrophy vector with gene expression. Gene set enrichment analysis was done to identify significant terms enriched in MSA-relevant regions. *Abbreviations: HC, healthy controls; MSA, multiple system atrophy; SD, standard deviation; T1-w, T1-weighted*.

To extract morphological information from brainstem nuclei of interest, we performed deformation-based morphometry (DBM) on each subject’s T1-weighted image using the Computational Anatomy Toolbox (CAT12; r1742) in Statistical Parametric Mapping software (SPM12)^18^ and MATLAB (version R2019b). The Brainstem Navigator toolkit (v0.9)^19,20^ was then applied to each participant’s MNI-registered DBM map to extract the extent of deformation within three regions involved in MSA pathophysiology, namely the SN, inferior olive, and superior olive.

To characterize atrophy at a finer granularity, vertex-wise surface-based analysis of cortical thickness in FreeSurfer was performed. A 15-mm smoothing kernel was applied to the surfaces.

Quality control was performed visually by an experienced neuroradiologist (L.C.). Bilateral regions were analyzed separately.

### Processing of atrophy maps

Morphological values were harmonized using ComBat, a correction tool that removes scanner-related variability while preserving biological effects of interest.^21^ A W-scoring approach was applied to the ComBat-corrected values to remove the effects of age and sex and derive deviations from what is expected for age and sex based on regressions generated within the control group.^10–12^ A negative W-score indicates atrophy, whereas a positive W-score indicates expansion, accounting for normal aging and sex effects.

### Regional gene expression extraction

To characterize the gene expression patterns associated with regions showing atrophy in MSA, we performed an imaging transcriptomics approach.^8^ We used abagen (v0.1.3)^22^ to extract the regional expression values of >20,000 genes from the Allen Human Brain Atlas (AHBA)^23^ in the regions showing atrophy. The AHBA reports transcriptional activity measurements for almost the entire genome, quantified in 3702 tissue samples from six postmortem brains. Two cerebellar lobules which did not match with any AHBA region (vermis and vermis VII) were discarded, resulting in 26 cerebellar regions. The main analysis focused on deep brain regions due to atrophy being predominant in these areas using a region-by-gene expression matrix of 15,611 genes by 50 regions. Since only two brains had available right hemisphere gene expression values, measurements from the left hemisphere were mirrored onto the right hemisphere to ensure whole-brain transcriptomic coverage. These gene expression values were used as predictors for the partial least squares regression.

### Partial least squares regression

Partial least squares (PLS) regression was used to identify gene expression patterns associated with deep brain atrophy in MSA. PLS regression is a multivariate approach that identifies latent variables that explain maximal covariance between two matrices, here a matrix of atrophy **X** (65 patients after quality control by 50 regions) and a matrix of regional gene expression **Y** (15,611 genes by 50 regions). The matrices **X** and **Y** were multiplied to generate a correlation matrix, which was then subjected to singular value decomposition. The statistical significance of the latent variables was assessed by comparing the empirical covariance explained by each latent variable to the covariance explained by 10,000 null models where atrophy was randomly permuted between regions (random null models). Given that the brain is characterized by a high degree of spatial autocorrelation between brain regions, we also tested the significance of latent variables against 10,000 spatially-constrained null models generated with BrainSMASH.^24^ A latent variable was considered significant when fewer than 5% of the null models explained more covariance than the original latent variable (P<0.05).

To identify the genes most robustly associated with each latent variable, we randomly shuffled the rows of **X** and **Y** with bootstrap resampling and repeated the PLS regression. This was repeated 5000 times to obtain bootstrap ratio weights from the generated null distribution. The ranked gene lists were used as inputs for gene set enrichment analysis.

### Gene set enrichment analysis

To identify the biological processes, cellular components, and human diseases gene terms overexpressed in association with atrophy in MSA, we performed gene set enrichment analysis (GSEA) in WebGestalt 2024.^25^ Gene Ontology terms^26^ were used for biological processes and cellular components, and DisGeNET terms^27^ for human diseases.

GSEA assessed whether negatively-weighted genes (associated with atrophy) were found more frequently within certain gene terms.^25^ Only gene terms with a minimum of 20 and maximum of 2000 genes were considered. Significance was determined using 1000 random permutations and false discovery rate (FDR) correction for multiple comparisons. To ensure that our results were independent of the enrichment platform used, we repeated the GSEA using PANTHER (version 19.0, 20240619).^28^

To identify which terms were specifically overexpressed in each MSA subgroup, we ran additional sub-analyses in each subgroup separately.

### Cell type analysis

Two transcriptomic-based approaches were performed to identify the cell types whose genes were significantly expressed in relation to atrophy. First, we used a GSEA as described above using WebGestalt 2024^25^ and the Human Cell Landscape,^29^ which analyzes cells from >50 human tissues. Second, using single-cell RNA sequencing performed specifically on cortical samples,^30^ we extracted the gene expression associated with oligodendrocytes, oligodendrocyte progenitor cells, microglia, astrocytes, excitatory neurons, inhibitory neurons, and endothelial cells, from deep brain regions.^13^ For each cell type, we computed the regional average expression across all genes and then calculated Pearson’s correlations between the average gene expression and atrophy maps. Correlations were corrected for 7 multiple comparisons with Bonferroni (P<0.007), tested against 10,000 spatial null models, and resulting P values were further FDR-corrected.^24^

### Specificity analysis: comparison with PD

To test whether the gene enrichment patterns in MSA were disease-specific, we replicated the same analysis on a sample of 57 patients with PD and 57 age- and sex-matched healthy controls recruited through the Quebec Parkinson Network (QPN;), and scanned on a 3T Siemens PRISMA machine at the Montreal Neurological Institute-MNI, Montreal.^31^ These participants were age- and sex-matched with MSA patients.

### Neurotransmitter mapping

We investigated whether atrophy in MSA occurred in regions overexpressing certain neurochemical systems. We parcellated and z-scored the regional density maps of 19 receptors, transporters, and binding sites to quantify the density of dopamine (D1, D2, dopamine transporter [DAT]), serotonin (5-HT_1A_, 5-HT_1B_, 5-HT_2A_, 5-HT_4_, 5-HT_6_, serotonin transporter [5-HTT]), noradrenaline (noradrenaline transporter [NET]), acetylcholine (α4β2, vesicular acetylcholine transporter [VAChT], M1), GABA (GABA_A_/BZ), glutamate (mGluR5, NMDA), histamine (H3), endocannabinoids (CB1), and opioids (μ)^13^ in the same regions as atrophy using *neuromaps* (details and references in Appendix).^32^ Cerebellar regions were excluded since PET images were normalized to the cerebellum for most tracers. Spearman’s correlation was calculated between each neurochemical regional density map and atrophy W-scores. Correlations were corrected for 19 multiple comparisons with Bonferroni (P<0.003), tested against 10,000 spatial null models, and resulting P values were further FDR-corrected.

### Statistical analysis

Differences in sex proportion and age were compared between controls and MSA patients using chi-squared test and Kruskal-Wallis test, respectively. One-sample t-tests were conducted on the atrophy W-scores to characterize the regions where MSA patients differed from controls and corrected with FDR. Effects of laterality on the atrophy W-scores were investigated in MSA using paired t-tests and FDR correction. Regarding the vertex-wise surface analysis, general linear modeling was done at each vertex to test the significance of a group effect on cortical thickness while controlling for age and sex; clusters of reduced thickness were considered significant using cluster- and voxel-level P values of 0.05 and 0.0001 and Monte-Carlo simulation. Region and vertex-wise comparisons were also performed between the MSAp and MSAc subgroups. Specifics on the imaging and statistical methods have been described in previous studies.^10–13^

## Results

### Participants

A total of 69 MSA patients and 190 controls were included. Of these, 13 (4 MSA, 9 controls) failed image processing or quality control, resulting in 246 participants, namely 65 MSA patients and 181 controls. The MSA group included 34 MSAp, 22 MSAc, and 9 mixed. MSA patients did not differ from controls in age (MSA: 61.7 ± 8.0, controls 63.2 ± 7.7; P=0.19) or sex (MSA: 41.5% female, controls 49.7% female; P=0.32). Average disease duration was 4.0 ± 2.0 years and the Unified Parkinson’s Disease Rating Scale (UPDRS) motor score was 33.3 ± 21.2 (Table 1).

**TABLE 1.** Demographic and clinical characteristics of participants.

|  | Controls | MSA |  |  |  | P value |
| --- | --- | --- | --- | --- | --- | --- |
|  |  | Total | MSAp | MSAc | MSA mixed |  |
| **n** | 181 | 65 | 34 | 22 | 9 |  |
| **Age (years)** | 63.2 ± 7.7 | 61.7 ± 8.0 | 63.3 ± 8.5 | 59.1 ± 6.2 | 62.4 ± 9.1 | 0.188 |
| **Females, n (%)** | 90 (49.7%) | 27 (41.5%) | 15 (44.1%) | 8 (36.4%) | 4 (44.4%) | 0.323 |
| **Disease duration (years)** | -- | 4.0 ± 2.0 | 3.8 ± 1.9 | 4.3 ± 2.2 | 3.9 ± 2.7 | 0.792 |
| **UPDRS III scores<sup>1</sup>** | -- | 33.3 ± 21.2 | 36.3 ± 21.4 | 31.7 ± 20.0 | 30.0 ± 23.3 | 0.711 |

Differences in sex proportion and age were compared between controls and MSA patients using chi-squared test and Kruskal-Wallis test, respectively. Differences in disease duration and UPDRS part III scores were compared between MSA groups using Kruskal-Wallis test.
<sup>1</sup> Higher values indicate worse motor symptoms, with 108 being the maximum score.
*Abbreviations: MSA, multiple system atrophy; MSAp, parkinsonian variant of MSA; MSAc, cerebellar variant of MSA; UPDRS, Unified Parkinson's Disease Rating Scale.*

### MSA patients show subcortical and cortical brain atrophy

MSA patients had severe deep brain atrophy compared to controls, particularly in the cerebellar white matter and cortex, pons, putamen, superior and inferior olive, and SN, bilaterally (all P_FDR_<0.0001, Table S3). There was no asymmetry in atrophy but for the inferior olive (P_FDR_<0.0001). Cortical atrophy was seen in 46 (68%) regions, particularly in the frontal and parietal cortices. The most atrophic regions were the bilateral precentral and caudal middle frontal cortices, the right pars opercularis and supramarginal cortices, the left precuneus, inferior parietal, paracentral, and rostral middle frontal cortices (P_FDR_<0.0001, Fig 2, Table S3). There was no asymmetry in cortical atrophy. To characterize cortical changes at a finer granularity, we performed vertex-wise analysis on the cortical surface and found that cortical thinning occurred at a finer resolution than parcel-based resolution and was relatively symmetrical and prominent within the precentral gyrus (Fig S1 and Table S4).

**FIGURE 2.**
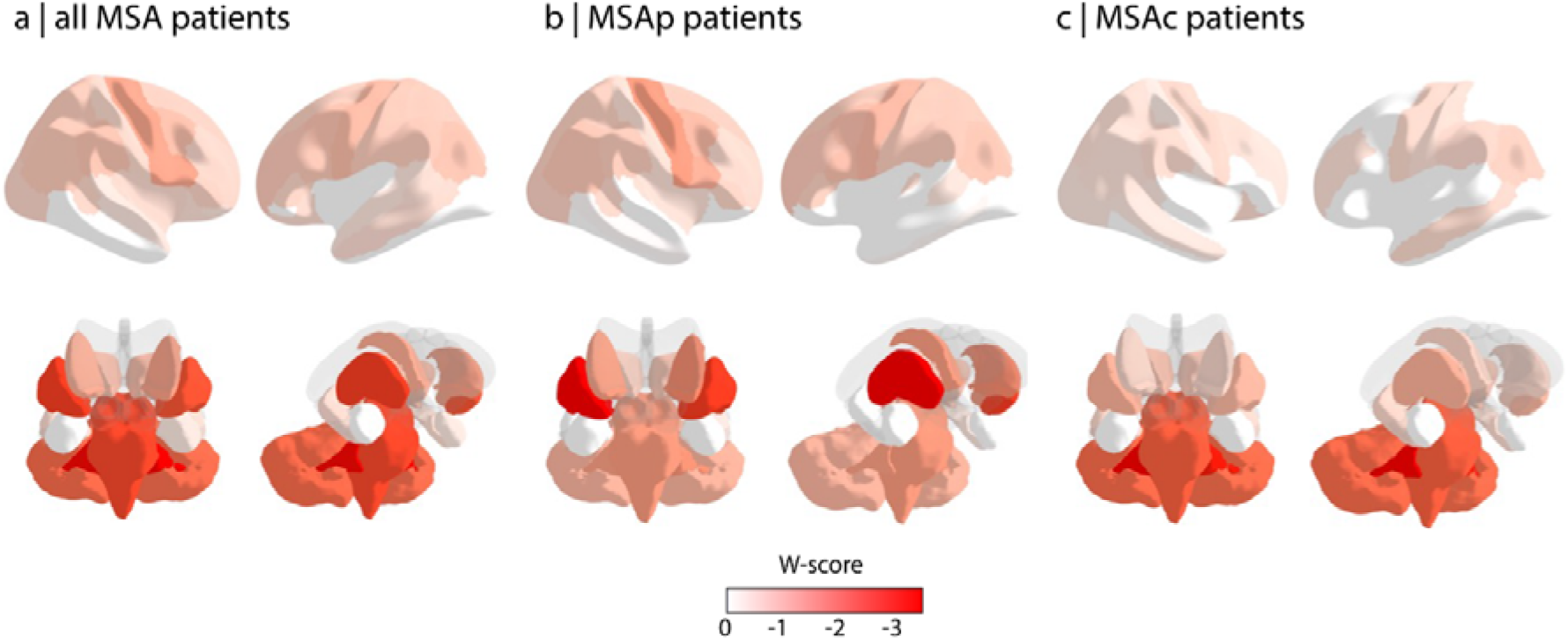
Pattern of atrophy in MSA. Regional W-scores associated with cortical (first row, left and right sagittal lateral views) and deep brain regions (second row, coronal and right oblique coronal views) were projected onto a brain volume in all MSA patients (a), those with MSAp (b), and those with MSAc (c). Red shades represent lower W-scores relative to controls, signifying greater atrophy; white shades indicate no significant atrophy. Only regions with W-scores significantly different from a null distribution are represented. Of note, the same color scale was used to depict atrophy in cortical and deep brain regions to maintain comparability of the magnitude of atrophy between the two sets of regions. *Abbreviations: MSA, multiple system atrophy; MSAp, parkinsonian variant of MSA; MSAc; cerebellar variant of MSA*.

When comparing MSAc and MSAp groups, as expected, MSAp patients had more atrophy in the putamen (P_FDR_<0.0001) and caudate (left: P_FDR_=0.006, right: P_FDR_=0.04, Table S5). Compared to MSAp patients, MSAc ones had greater atrophy in the cerebellar white matter (P_FDR_<0.0001) and cortex (P_FDR_<0.0001), followed by the pons (P_FDR_<0.0001), midbrain (P_FDR_<0.02), superior cerebellar peduncles (P_FDR_=0.02), and superior olive (P_FDR_≤0.008). The SN, inferior olive, and cortical regions did not differ between MSAp and MSAc patients (Table S5).

### The brain’s gene expression spatial distribution predicts atrophy in MSA

We next investigated whether deep brain atrophy in MSA related to the brain’s spatial distribution of gene expression. Of the 5 gene expression latent variables returned, two showed significance and explained 28% of the covariance in gene-atrophy compared to random (LV3: 19.8% of covariance explained vs. 11.3% in null models, P=0.03; LV5: 7.6% vs. 3.7%, P=0.02) and spatial null models (LV3: 12.1%, P=0.04; LV5: 3.7%, P=0.01; Fig 3a). For both variables, regions with lower W-scores (more atrophy) had more negative weights (LV3: r=0.44, P<0.001; LV5: r=0.28, P=0.05), meaning that negatively-weighted genes were more expressed in regions with greater atrophy (Fig 3b, 3d; Fig 4). This suggests that the brain’s gene expression pattern is associated with the pattern deep brain atrophy in MSA.

**FIGURE 3.**
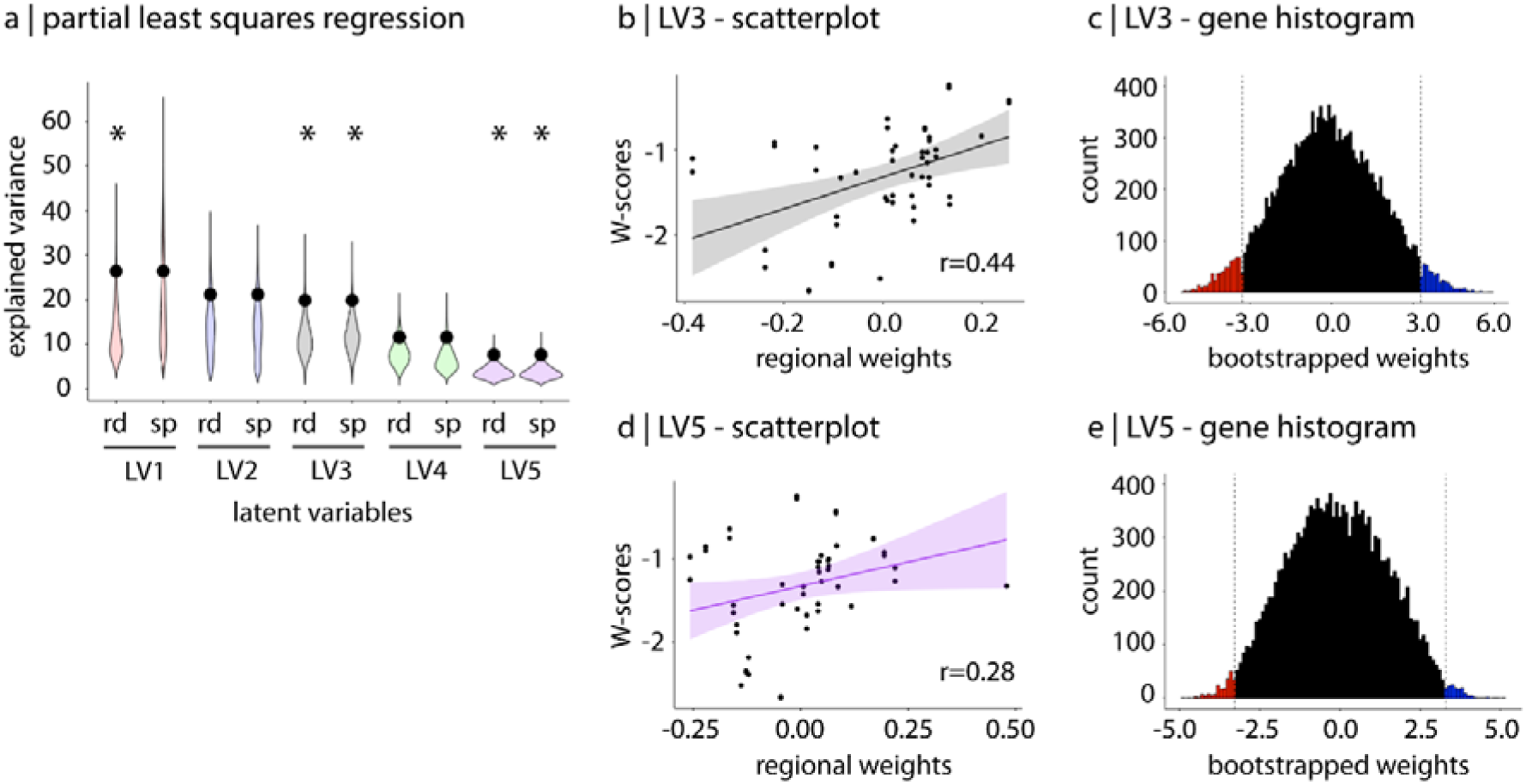
Patterns of gene expression in MSA. (a) Results of the partial least squares analysis on deep brain regions with violin plots representing the percentage of covariance in deep brain regions W-scores explained by gene expression (y-axis); the black dots indicate the empirical covariance, and the asterisks indicate the significant latent variables (LV) against random (rd) and spatial (sp) null models (x-axis). (b, d) Scatterplots of the correlation between W-scores associated with deep brain regions (x-axis) and regional weights of LV3 and LV5 (y axis). (c, e) Histograms of the genes’ bootstrapped weights on LV3 and LV5. Bootstrap weights represented in red are robustly associated with regions with greater atrophy (below −3.29). *Abbreviations: LV, latent variable; rd, random null models; sp, spatial null models*.

**FIGURE 4.**
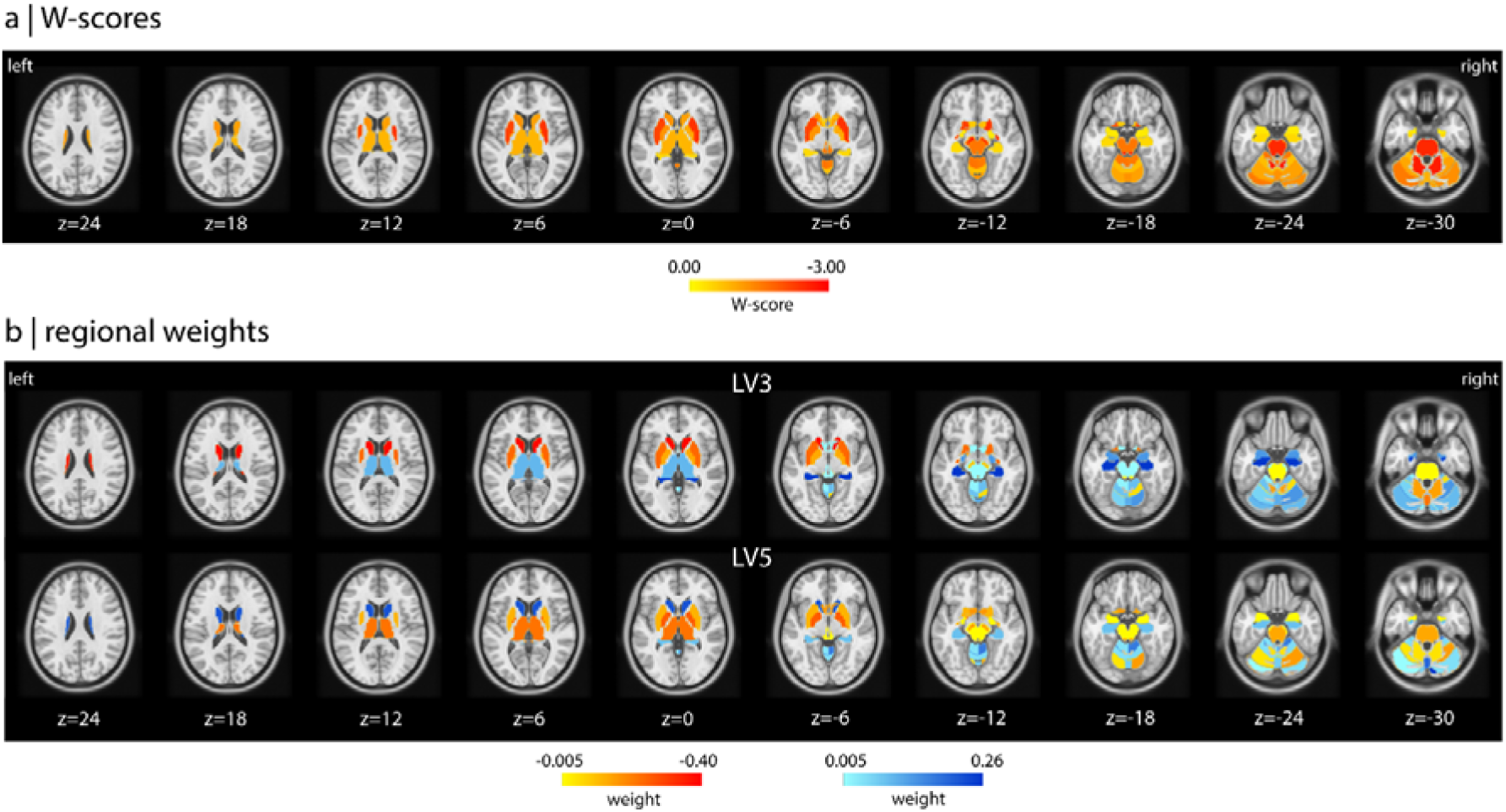
Brain maps of W-scores associated with deep brain regions and regional weights of the significant latent variables of the PLS. Brain maps of W-scores associated with deep brain regions in MSA patients (a) and spatial distribution of the regional weights associated with LV3 and LV5 (b). Coordinates along the z axis in the Montreal Neurological Institute (MNI) template are given for each slice level. *Abbreviations: LV, latent variable; PLS, partial least squares*.

### Brain atrophy is associated with mitochondrial function

Next, we performed GSEA to investigate the biological processes, cellular components, human disease terms, enriched in the genes most strongly associated with brain atrophy in MSA. When assessing the genes negatively weighted on LV3, 652 (4.2%) genes were associated with atrophy after bootstrapping (Fig 3c, 3e). In terms of biological processes, atrophic regions were enriched for processes related to the mitochondrial respiratory chain, with the most significant terms being proton transmembrane transport, complex I assembly, electron transport chain, mitochondrial transport, mitochondrion organization (all P_FDR_<0.0001), energy derivation by oxidation of organic compounds (P_FDR_<0.001), and nucleoside triphosphate metabolic process (P_FDR_<0.001) (Table 2, Fig 5). Regarding cellular components, regions showing atrophy were enriched for genes associated with the mitochondrial protein-containing complex, respirasome, oxidoreductase complex, mitochondrial inner membrane, and myelin sheath (all P_FDR_<0.0001). In terms of human diseases, atrophic regions overexpressed terms related to mitochondrial diseases (Fig 5, Table 2). Similar findings were obtained when performing GSEA using the PANTHER gene enrichment platform (Table S6).

**FIGURE 5.**
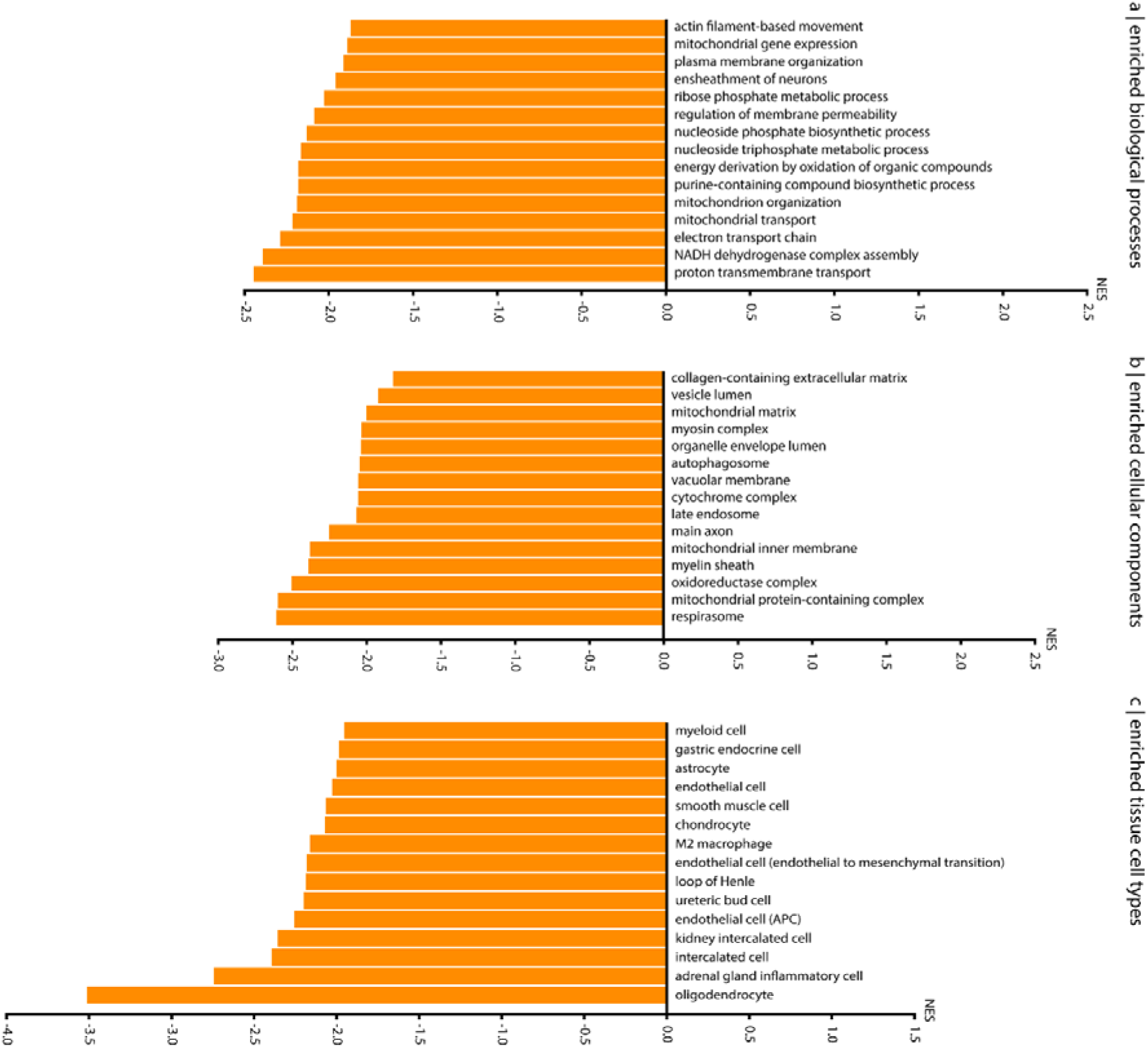
Gene set enrichment analysis in the whole MSA sample. Significant biological processes (a), cellular components (b), and tissue cell types (c) that were enriched in the negatively-weighted genes associated with deep brain atrophy in MSA. Terms are ranked from right to left based on the normalized enrichment score; all enriched terms were significant after FDR correction. Results were obtained with WebGestalt 2024.^25^ Only the top 15 terms are shown for visualization purposes. *Abbreviations: FDR, false discovery rate; NES, normalized enrichment score*.

**TABLE 2.** Biological processes, cellular components, and human disease terms enriched in the genes related to brain atrophy in MSA.

| Gene Set | Description | Size | Leading Edge Number | Enrichment score | Normalized enrichment score | P value | P<sub>FDR</sub> value |
| --- | --- | --- | --- | --- | --- | --- | --- |
| **Biological processes** |  |  |  |  |  |  |  |
| GO:1902600 | Proton transmembrane transport | 114 | 74 | -0.542 | -2.417 | <0.0001 | <0.0001 |
| GO:0010257 | NADH dehydrogenase complex assembly | 54 | 42 | -0.608 | -2.359 | <0.0001 | <0.0001 |
| GO:0022900 | Electron transport chain | 146 | 76 | -0.486 | -2.249 | <0.0001 | <0.0001 |
| GO:0006839 | Mitochondrial transport | 161 | 103 | -0.468 | -2.192 | <0.0001 | <0.0001 |
| GO:0007005 | Mitochondrion organization | 439 | 227 | -0.416 | -2.181 | <0.0001 | <0.0001 |
| GO:0015980 | Energy derivation by oxidation of organic compounds | 280 | 137 | -0.430 | -2.167 | <0.0001 | <0.0001 |
| GO:0009141 | Nucleoside triphosphate metabolic process | 223 | 128 | -0.440 | -2.161 | <0.0001 | <0.0001 |
| GO:0072522 | Purine-containing compound biosynthetic process | 216 | 126 | -0.444 | -2.156 | <0.0001 | <0.0001 |
| GO:1901293 | Nucleoside phosphate biosynthetic process | 240 | 129 | -0.427 | -2.099 | <0.0001 | 0.001 |
| GO:0090559 | Regulation of membrane permeability | 58 | 28 | -0.521 | -2.056 | <0.0001 | 0.002 |
| **Cellular components** |  |  |  |  |  |  |  |
| GO:0098798 | Mitochondrial protein-containing complex | 272 | 170 | -0.554 | -2.532 | <0.0001 | <0.0001 |
| GO:0070469 | Respirasome | 86 | 58 | -0.621 | -2.478 | <0.0001 | <0.0001 |
| GO:1990204 | Oxidoreductase complex | 107 | 66 | -0.575 | -2.375 | <0.0001 | <0.0001 |
| GO:0005743 | Mitochondrial inner membrane | 430 | 228 | -0.494 | -2.334 | <0.0001 | <0.0001 |
| GO:0043209 | Myelin sheath | 38 | 16 | -0.641 | -2.195 | <0.0001 | <0.0001 |
| GO:0031970 | Organelle envelope lumen | 84 | 38 | -0.544 | -2.160 | <0.0001 | <0.0001 |
| GO:0070069 | Cytochrome complex | 33 | 21 | -0.610 | -2.030 | <0.0001 | 0.0001 |
| GO:0005774 | Vacuolar membrane | 402 | 172 | -0.430 | -2.015 | <0.0001 | 0.0002 |
| GO:0005759 | Mitochondrial matrix | 451 | 196 | -0.420 | -1.992 | <0.0001 | 0.0004 |
| GO:0016459 | Myosin complex | 41 | 23 | -0.567 | -1.989 | <0.0001 | 0.0004 |
| **Human diseases** |  |  |  |  |  |  |  |
| umls:C2936907 | NADH:Q(1) oxidoreductase deficiency | 20 | 17 | -0.764 | -2.343 | <0.0001 | <0.0001 |
| umls:C1838979 | Mitochondrial complex deficiency | 21 | 17 | -0.737 | -2.278 | <0.0001 | <0.0001 |
| umls:C0022548 | Keloid | 26 | 16 | -0.607 | -1.989 | <0.0001 | 0.010 |
| umls:C0029456 | Osteoporosis | 48 | 27 | -0.507 | -1.939 | <0.0001 | 0.018 |
| umls:C0007959 | Charcot-Marie-Tooth disease | 26 | 16 | -0.572 | -1.930 | <0.0001 | 0.015 |
| umls:C0023264 | Leigh disease | 23 | 18 | -0.589 | -1.888 | <0.0001 | 0.018 |
| umls:C0026640 | Mouth neoplasms | 31 | 13 | -0.545 | -1.880 | 0.00170 | 0.017 |
| umls:C0007134 | Renal cell carcinoma | 106 | 37 | -0.412 | -1.808 | <0.0001 | 0.031 |
| umls:C0029408 | Degenerative polyarthritis | 81 | 34 | -0.426 | -1.795 | <0.0001 | 0.032 |
| umls:C0878544 | Cardiomyopathies | 72 | 32 | -0.420 | -1.745 | <0.0001 | 0.037 |

The significant biological processes, cellular components, and disease terms enriched in the genes associated with deep brain atrophy in MSA are listed ranked based on the normalized enrichment score. Only the top 10 most significant terms results associated with negatively-weighted genes (i.e., genes more expressed in regions with greater atrophy) are reported. Results were obtained with WebGestalt 2024.<sup>25</sup>
*Abbreviations: FDR, false discovery rate; GO, gene ontology; NADH, Nicotinamide adenine dinucleotide phosphate.*

When assessing the enrichment pattern for the other significant latent variable (LV5), 219 (1.4%) genes were associated with greater atrophy (Fig 3e). GSEA revealed similar findings as in LV3, with enrichment of processes related to energy production and metabolism. When performing additional sub-analyses on the MSAp and MSAc subgroups separately, we also identified terms related to mitochondrial functions in both subgroups (Table S7).

Taken together, this results demonstrate that atrophy in MSA patients occurs preferentially in regions overexpressing genes associated with mitochondrial processes.

### Brain atrophy relates to oligodendrocytes

Using GSEA, cell type-related genes were overexpressed in relation to atrophy in MSA, including brain-related oligodendrocytes and endothelial cells (both P_FDR_<0.001) (Fig 5). Similarly, using single-cell RNA sequencing specific to neural cells, we found a negative correlation between W-scores in deep brain regions and gene expression of oligodendrocytes (r=-0.39, P=0.0006; P_FDR_ spatial=0.002), meaning a higher gene expression of these cell types in regions with greater atrophy (Table S8). These results support that atrophic regions overexpress gene expression profiles related to oligodendrocytes.

### Specificity analysis: comparison with PD

To assess the specificity of these findings to MSA, we repeated the PLS regression in 57 PD patients age- and sex-matched to the MSA patients. We found two significant latent variables explaining in total 20.0% of the covariance between gene expression and atrophy in PD. Regions associated with negatively-weighted genes (atrophy) in PD were enriched for synaptic functions. In terms of cellular components, terms related to synapses were significant, including postsynaptic specialization, neuron spine, neuron-to-neuron synapse, GABA-ergic synapse (Fig S2). There was no significant enrichment of cell types, including oligodendrocytes. These findings suggest that atrophy patterns in MSA and PD are supported by distinct gene expression profiles in MSA and PD, with mitochondrial functions and oligodendrocytes being specific to MSA.

### Brain atrophy maps onto specific neurotransmitter systems

We further tested whether the pattern of cortical and deep brain regions atrophy (cerebellum excluded) in MSA mapped onto specific neurotransmitter systems. Atrophic regions had lower density of serotonin receptors, namely 5-HT_1A_ (r=0.75, P<0.0001; P_FDR_ spatial<0.0001) and 5-HT_2A_ (r=0.36, P=0.001; P_FDR_ spatial =0.046), and GABA_A_/BZ receptors (r=0.42, P<0.0001; P_FDR_ spatial=0.046), as well as a higher density of α4β2 acetylcholine (r=-0.61, P<0.0001; P_FDR_ spatial <0.0001) and NET noradrenaline transporters (r=-0.36, P=0.001; P_FDR_ spatial=0.046) (Fig 6, Table S9).

**FIGURE 6.**
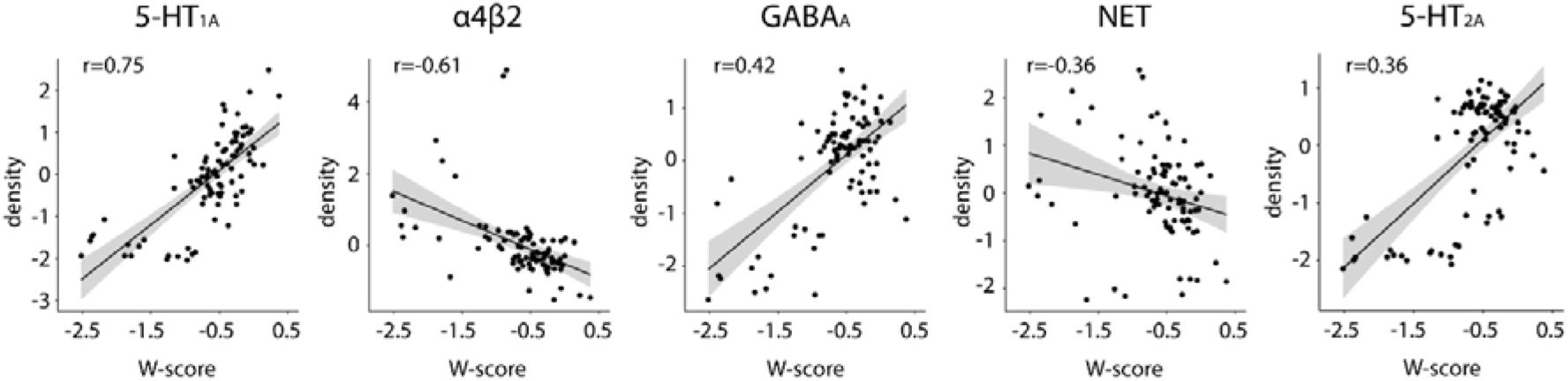
Neurotransmitter mapping. Scatterplots with linear regression lines between regional W-scores and density values of receptors in cortical and deep brain regions. Only significant correlations are plotted. A correlation was considered significant when the P value was below the Bonferroni-corrected threshold of P<0.003. *Abbreviations: FDR, false discovery rate*

## Discussion

Our study showed that the pattern of atrophy in MSA patients involved cortical and deep regions, being more predominant in the putamen, cerebellar white matter, and pons. We found that the regions showing atrophy demonstrate distinct gene expression profiles, related to mitochondrial functions and oligodendrocytes. Moreover, regions with greater atrophy colocalized with areas showing greater distribution of acetylcholine and noradrenaline receptors/transporters and lower distribution of serotonin and GABA receptors. This study highlights some of the biological and neurochemical underpinnings of selective vulnerability of brain regions to neurodegeneration in MSA.

As expected, we found that atrophy in MSA was more severe in deep brain regions, mainly involving the putamen in the parkinsonian variant, and the cerebellum, pons, and olive in the cerebellar subtype.^6,7^ In addition, atrophy was also present in specific cortical areas, notably in the bilateral precentral and caudal middle frontal cortices, the right pars opercularis region, and the left precuneus, in line with previous studies.^33^ The involvement of the precentral cortices, more prominent in MSAp than MSAc, may reflect extrapyramidal and corticospinal involvement. Subcortical degeneration may alter cortico-subcortical loops, potentially leading to structural reorganization in connected cortical structures.^33^ Moreover, MSA patients often present with pyramidal signs,^1^ reflecting corticospinal tract damage, as reported in neuropathological ^34^ and MRI studies.^35^

Using imaging transcriptomics, we found that atrophic regions in MSA overexpressed genes involved in mitochondrial assembly and functioning, suggesting that mitochondrial dysfunction represents an important factor of selective vulnerability to neurodegeneration. Mitochondria play a crucial role in several neurodegenerative diseases.^36^ As for PD,^37^ there is evidence of mitochondrial dysfunction in the pathogenesis of MSA based on genome-wide association studies,^4^ postmortem studies,^37,38^ and cell^39,40^ and animal models.^36^ Mutations in the COQ2 gene, encoding an enzyme involved in Coenzyme Q10 (CoQ10) biosynthesis, have been linked to familial and sporadic cases of MSA.^4^ CoQ10 is found in the inner mitochondrial membrane and is involved in the transfer of electrons from complexes I/II to complex III, thus playing an essential role in the functioning of the respiratory chain and the ATP production. Impaired CoQ10 activity increases vulnerability to oxidative stress.^36,37^ Autopsy studies have shown reduced CoQ10 levels in the cerebrospinal fluid ^41^ and blood ^42^ of MSA patients as well as in cerebellar^37,38^ and motor cortex postmortem samples.^38^ One study that investigated mitochondrial function within dopaminergic neurons derived from induced pluripotent stem cells (iPSCs) of MSA patients provided evidence for impaired respiratory chain activity and up-regulation of CoQ10 biosynthesis, the latter indicating possible compensatory mechanism.^39^ Another study found that exposure to oxidative stress of neural progenitor cells derived from MSA patient iPSCs triggered excessive generation of reactive oxygen species and cell damage.^40^ Building on these findings, our study reinforces the evidence of mitochondrial dysfunction in MSA using brain atrophy quantified from T1-weighted scans. Our results resemble those seen in other synucleinopathies, namely PD and iRBD, showing that cortical brain atrophy occurs in regions overexpressing genes involved in mitochondrial function or synaptic functioning.^10,12^ However, when repeating the same analyses on deep brain regions in PD, we found an overexpression of synapse-related genes, as in previous studies.^11^ Interestingly, no mitochondria-related terms were associated with deep brain regions in PD. These findings suggest that atrophy patterns exhibit distinct gene expression profiles in MSA and PD, and that the observed relationship between deep brain atrophy pattern and mitochondrial function and oligodendrocytes may be specific to MSA.

Another key finding of our study is the overexpression of genes involved in oligodendrocytes in regions showing greater atrophy in MSA patients, but not in PD, aligning with the fact that oligodendrocytes are the primary target of α-synuclein aggregates in MSA.^2,3^ This finding mirrors the association with genes enriched for mitochondrial functions given the energetic need of oligodendrocytes, especially for the process of myelination, which is particularly high. Whether mitochondrial dysfunction is a primary mechanism underlying oligodendropathy or is secondary to other pathological processes, especially α-synuclein accumulation, remains to be determined. Exploring the interplay between oligodendroglia and neurons will help to understand the association between the density of oligodendrocytes and brain atrophy observed in our study. Indeed, α-synuclein-mediated oligodendroglial pathology contributes to neuronal damage, as supported by the positive correlation between neuronal loss and glial cytoplasmic inclusion density.^3^ Oligodendrocytes are crucial not only in the formation of the myelin sheath, but also for providing trophic support to neurons. Factors released by oligodendroglial precursors and mature oligodendrocytes are essential for neuronal survival. Consequently, the deficiency of oligodendroglia-derived neurotrophic factors resulting from oligodendroglial impairment may account for the neuronal loss.^36^ In sum, our results support the theory that neurodegeneration in MSA may derive from neuronal energy failure and lack of trophic support from oligodendroglia.

Regions most vulnerable to atrophy in MSA colocalized with areas with higher distribution of NET (noradrenaline) and α4β2 (acetylcholine), and lower distribution of serotonin (5-HT_1A_/5-HT_2A_) and GABA receptors. These findings support the involvement of multi-neurochemical systems related to neuronal cell loss across several neurotransmitter projection systems, as demonstrated by postmortem and PET studies.^43^ To date, only one study has applied the spatial mapping approach in MSA to investigate the neurochemical underpinnings of iron deposition patterns, demonstrating that regions with higher cortical iron content overlapped with areas of higher density of noradrenaline and acetylcholine receptors.^44^ Central autonomic pathways, crucial for autonomic cardiovascular and respiratory control, are impaired in MSA as a result of a central noradrenergic deficiency.^45^ This noradrenergic denervation is reflected by a decrease of cerebrospinal fluid levels of noradrenaline, coupled with reduced noradrenaline density in the frontal cortex and putamen on post-mortem tissue analysis.^46^ The locus coeruleus (LC) is the main source of noradrenaline in the brain. Noradrenergic neurons from this area project widely throughout the brain and spinal cord, and are involved in arousal, attention, and stress responses.^47^ It is possible that central noradrenergic deficiency, possibly related to a lesion of the LC, contributes to baroreflex dysfunction that in turn induces the orthostatic hypotension observed in MSA. The LC is also known to project to the nucleus of the solitary tract, where all baroreceptor afferents initially synapse in the brain, and to the rostral ventrolateral medulla, a major source of descending projections to sympathetic pre-ganglionic neurons and critical area for the tonic maintenance of sympathetic vasomotor tone and blood pressure control. The association between atrophy and higher cholinergic receptor density shown here aligns with postmortem studies demonstrating severe depletion of cholinergic neurons in the pedunculopontine and laterodorsal tegmental nuclei and LC.^47^ Similarly, using 1-[^11^C]Methylpiperidin-4-yl propionate, a PET radiotracer targeting acetylcholinesterase, a cholinergic activity reduction was shown in MSAp patients, similar to PD in the cortex, but more pronounced in subcortical regions, potentially accounting for the gait and cognitive disturbances observed in MSAp.^49^ Furthermore, there is evidence of serotonergic neuronal loss in the caudal brainstem raphe nucleus in MSA, as shown by neuropathological^43^ and PET imaging studies reporting decreased cortical serotonin transporter binding.^50^ Depletion of serotonergic neurons may contribute to impaired autonomic and respiratory control.

Our study has limitations. First, there was no pathological confirmation of diagnoses. Instead, we only included patients meeting criteria for clinically established MSA, which is currently the highest level of diagnostic certainty. Second, although the sample size was relatively small, given the rarity of the disease, it represents a significant group of patients for exploring brain abnormalities. Third, due to the retrospective nature of this cohort, we had access to limited clinical data, restricting the investigation of correlations between clinical features and atrophy. Furthermore, the gene expression data came from six healthy donors with varying ages and medical histories. Future studies should use transcriptomic atlases from a larger number of controls closely matched to the MSA population.

To conclude, we showed that atrophy in MSA occurs mainly in deep brain regions overexpressing genes involved in mitochondrial functioning and oligodendrocytes, and maps onto specific neurochemical systems. Based solely on morphological measurements derived from MRI scans, we were able to identify biological correlates of brain atrophy in MSA. Future studies should investigate whether the neural bases of clinical features associated with MSA map to differential transcriptomic and neurochemical systems.

## List of abbreviations

DBM: deformation-based morphometry
FDR: false discovery rate
GO: gene ontology
iRBD: isolated rapid eye movement sleep behaviour disorder
LV: latent variable
MSA: multiple system atrophy
MSAp: parkinsonian variant of MSA
MSAc: cerebellar variant of MSA
PD: Parkinson’s disease
PLS: partial least squares.

## Acknowledgement statement

This work was supported by grants from Agence Nationale de la Recherche (grant numbers ANR-11-INBS-0006 [France Life Imaging], ANR-11-INBS-0011 [NeurATRIS, Investissements d’Avenir], ANR-19-P3IA-0001 [PRAIRIE 3IA Institute], and ANR-10-IAIHU-06 [IHU–Paris Institute of Neurosciences]), the European Union (EU) Framework Project 6–GENEPARK: Genomic Biomarkers for Parkinson’s Disease, Action Line: LIFESCIHEALTH Life sciences, genomics and biotechnology for health (LSH-2005-1.2.2.2)—Development of Innovative methods for diagnosis of nervous system disorders, the Programme Hospitalier de Recherche Clinique [grant Numbers: PHRC 2007-A00169-44 (LRRK) and PHRC 2004 (BBBIPPS), Association France Parkinson, Ecole Neuroscience de Paris, Electricité de France (Fondation d’Entreprise EDF), Institut National de la Santé et de la Recherche Médicale, Fondation Thérèse and René Planiol pour l’étude du Cerveau, Société Française de Radiologie (SFR) / Collège des Enseignants en Radiologie de France (CERF), and Société Française de Neuroradiologie (SFNR).

The work was also supported by Fonds de recherche du Quebec – Santé (FRQS), the Canadian Institutes of Health Research (CIHR) and the Healthy Brains for Healthy Lives(HBHL) program of the Canada First Research Excellence Fund.

## Author contributions

Lydia Chougar designed and conceptualized the study, collected and analyzed the data, drafted and revised the manuscript for intellectual content. Christina Tremblay, Aline Delva, Marie Filiatrault, Andrew Vo, Justine Y. Hansen, Asa Farahani, Parsa Khalafi, Charles-Etienne Castonguay, Guy Rouleau and Alain Dagher analyzed the data and revised the manuscript. Bratislav Misic assisted with the methodology and revised the manuscript. Jean-Christophe Corvol, Marie Vidailhet, Bertrand Degos, David Grabli, and Stéphane Lehéricy performed data collection and revised the manuscript. Shady Rahayel: designed and conceptualized the study, collected and analyzed the data, drafted and revised the manuscript for intellectual content

## Data availability

The data used in this study originate from local cohorts and clinical studies, which are governed by data-sharing agreements that strictly prohibit the distribution of individual-level data. These restrictions ensure the confidentiality and privacy of participant information in compliance with ethical and regulatory standards. However, group-level aggregate data, such as summary statistics or derived metrics, can be shared upon reasonable request to the corresponding authors.

## References

1. Wenning GK, Stankovic I, Vignatelli L, et al. The Movement Disorder Society Criteria for the Diagnosis of Multiple System Atrophy. Movement Disorders 2022;mds.29005.

2. Dickson DW. Parkinson’s Disease and Parkinsonism: Neuropathology [Internet]. Cold Spring Harb Perspect Med 2012;2(8)[cited 2019 Mar 16] Available from: https://www.ncbi.nlm.nih.gov/pmc/articles/PMC3405828/

3. Jellinger KA. Neuropathology of multiple system atrophy: new thoughts about pathogenesis. Mov. Disord. 2014;29(14):1720–1741.

4. Multiple-System Atrophy Research Collaboration. Mutations in COQ2 in familial and sporadic multiple-system atrophy. N Engl J Med 2013;369(3):233–244.

5. Sailer A, Scholz SW, Nalls MA, et al. A genome-wide association study in multiple system atrophy. Neurology 2016;87(15):1591–1598.

6. Chougar L, Pyatigorskaya N, Degos B, et al. The Role of Magnetic Resonance Imaging for the Diagnosis of Atypical Parkinsonism. Front. Neurol. 2020;11:665.

7. Chougar L, Pyatigorskaya N, Lehéricy S. Update on neuroimaging for categorization of Parkinson’s disease and atypical parkinsonism [Internet]. Current Opinion in Neurology 2021;Publish Ahead of Print[cited 2022 Mar 2] Available from: https://journals.lww.com/10.1097/WCO.0000000000000957

8. Arnatkeviciute A, Markello RD, Fulcher BD, et al. Toward Best Practices for Imaging Transcriptomics of the Human Brain. Biological Psychiatry 2023;93(5):391–404.

9. Thomas GEC, Leyland LA, Schrag A-E, et al. Brain iron deposition is linked with cognitive severity in Parkinson’s disease. J Neurol Neurosurg Psychiatry 2020;jnnp-2019–322042.

10. Vo A, Tremblay C, Rahayel S, et al. Network connectivity and local transcriptomic vulnerability underpin cortical atrophy progression in Parkinson’s disease. NeuroImage: Clin. 2023;40:103523.

11. Tremblay C, Rahayel S, Vo A, et al. Brain atrophy progression in Parkinson’s disease is shaped by connectivity and local vulnerability. Brain Commun 2021;3(4):fcab269.

12. Rahayel S, Tremblay C, Vo A, et al. Mitochondrial function-associated genes underlie cortical atrophy in prodromal synucleinopathies. Brain 2023;146(8):3301–3318.

13. Hansen JY, Shafiei G, Markello RD, et al. Mapping neurotransmitter systems to the structural and functional organization of the human neocortex. Nat. Neurosci. 2022;25(11):1569–1581.

14. Marek K, Chowdhury S, Siderowf A, et al. The Parkinson’s progression markers initiative (PPMI) – establishing a PD biomarker cohort. Annals of Clinical and Translational Neurology 2018;5(12):1460.

15. Iglesias JE, Van Leemput K, Bhatt P, et al. Bayesian segmentation of brainstem structures in MRI. Neuroimage 2015;113:184–195.

16. Faber J, Kügler D, Bahrami E, et al. *CerebNet*: A fast and reliable deep-learning pipeline for detailed cerebellum sub-segmentation. NeuroImage 2022;264:119703.

17. Schmahmann JD, Doyon J, McDonald D, et al. Three-dimensional MRI atlas of the human cerebellum in proportional stereotaxic space. Neuroimage 1999;10(3 Pt 1):233–260.

18. Gaser C, Dahnke R, Thompson PM, et al. CAT – A Computational Anatomy Toolbox for the Analysis of Structural MRI Data. bioRxiv 2023;2022.06.11.495736.

19. García_Gomar MG, Singh K, Cauzzo S, Bianciardi M. In vivo structural connectome of arousal and motor brainstem nuclei by 7 Tesla and 3 Tesla MRI. Hum. Brain Mapp. 2022;43(14):4397– 4421.

20. Singh K, Cauzzo S, García-Gomar MG, et al. Functional connectome of arousal and motor brainstem nuclei in living humans by 7 Tesla resting-state fMRI. NeuroImage 2022;249:118865.

21. Fortin J-P, Cullen N, Sheline YI, et al. Harmonization of cortical thickness measurements across scanners and sites. NeuroImage 2018;167:104–120.

22. Markello RD, Arnatkeviciute A, Poline J-B, et al. Standardizing workflows in imaging transcriptomics with the abagen toolbox [Internet]. eLife 2021;10 Available from: https://app.readcube.com/library/a2839d09-3508-4669-a1c5-2914e8010a23/item/79c2c9e0-69b6-428f-9123-2dd59cf22cdc

23. Hawrylycz MJ, Lein ES, Guillozet-Bongaarts AL, et al. An anatomically comprehensive atlas of the adult human brain transcriptome. Nature 2012;489(7416):391–399.

24. Burt JB, Helmer M, Shinn M, et al. Generative modeling of brain maps with spatial autocorrelation. NeuroImage 2020;220:117038.

25. Elizarraras JM, Liao Y, Shi Z, et al. WebGestalt 2024: faster gene set analysis and new support for metabolomics and multi-omics. Nucleic Acids Research 2024;52(W1):W415–W421.

26. Consortium TGO, Aleksander GCSA, Balhoff J, et al. The Gene Ontology knowledgebase in 2023. Genetics 2023;224(1):iyad031.

27. Piñero J, Ramírez-Anguita JM, Saüch-Pitarch J, et al. The DisGeNET knowledge platform for disease genomics: 2019 update. Nucleic Acids Research 2019;48(D1):D845.

28. Mi H, Muruganujan A, Huang X, et al. Protocol Update for Large-scale genome and gene function analysis with PANTHER Classification System (v.14.0). Nat Protoc 2019;14(3):703– 721.

29. Han X, Zhou Z, Fei L, et al. Construction of a human cell landscape at single-cell level. Nature 2020;581(7808):303–309.

30. Seidlitz J, Nadig A, Liu S, et al. Transcriptomic and cellular decoding of regional brain vulnerability to neurogenetic disorders. Nat Commun 2020;11(1):3358.

31. Gan-Or Z, Rao T, Leveille E, et al. The Quebec Parkinson Network: A Researcher-Patient Matching Platform and Multimodal Biorepository. J. Park.’s Dis. 2020;10(1):301–313.

32. Markello RD, Hansen JY, Liu Z-Q, et al. neuromaps: structural and functional interpretation of brain maps. Nat. Methods 2022;19(11):1472–1479.

33. Fiorenzato E, Weis L, Seppi K, et al. Brain structural profile of multiple system atrophy patients with cognitive impairment. J Neural Transm (Vienna) 2017;124(3):293–302.

34. Lin CR, Viswanathan A, Chen TX, et al. Clinicopathological correlates of pyramidal signs in multiple system atrophy. Ann Clin Transl Neurol 2022;9(7):988–994.

35. Ogawa T, Hatano T, Kamagata K, et al. White matter and nigral alterations in multiple system atrophy-parkinsonian type. npj Parkinsons Dis. 2021;7(1):1–12.

36. Compagnoni GM, Fonzo AD. Understanding the pathogenesis of multiple system atrophy: state of the art and future perspectives [Internet]. Acta Neuropathologica Communications 2019;7[cited 2024 Aug 9] Available from: https://www.ncbi.nlm.nih.gov/pmc/articles/PMC6624923/

37. Foti SC, Hargreaves I, Carrington S, et al. Cerebral mitochondrial electron transport chain dysfunction in multiple system atrophy and Parkinson’s disease. Sci Rep 2019;9(1):6559.

38. Hsiao J-HT, Purushothuman S, Jensen PH, et al. Reductions in COQ2 Expression Relate to Reduced ATP Levels in Multiple System Atrophy Brain. Front Neurosci 2019;13:1187.

39. Monzio Compagnoni G, Kleiner G, Samarani M, et al. Mitochondrial Dysregulation and Impaired Autophagy in iPSC-Derived Dopaminergic Neurons of Multiple System Atrophy. Stem Cell Reports 2018;11(5):1185–1198.

40. Herrera-Vaquero M, Heras-Garvin A, Krismer F, et al. Signs of early cellular dysfunction in multiple system atrophy. Neuropathology and Applied Neurobiology 2021;47(2):268–282.

41. Compta Y, Giraldo DM, Muñoz E, et al. Cerebrospinal fluid levels of coenzyme Q10 are reduced in multiple system atrophy. Parkinsonism Relat Disord 2018;46:16–23.

42. Kasai T, Tokuda T, Ohmichi T, et al. Serum Levels of Coenzyme Q10 in Patients with Multiple System Atrophy. PLoS One 2016;11(1):e0147574.

43. Benarroch EE, Schmeichel AM, Low PA, Parisi JE. Involvement of medullary serotonergic groups in multiple system atrophy. Ann Neurol 2004;55(3):418–422.

44. Yan S, Lu J, Duan B, et al. Quantitative susceptibility mapping of multiple system atrophy and Parkinson’s disease correlates with neurotransmitter reference maps. Neurobiol. Dis. 2024;198:106549.

45. Chelban V, Catereniuc D, Aftene D, et al. An update on MSA: premotor and non-motor features open a window of opportunities for early diagnosis and intervention. J Neurol 2020;267(9):2754–2770.

46. Goldstein DS, Sullivan P, Holmes C, et al. Differential abnormalities of cerebrospinal fluid dopaminergic versus noradrenergic indices in synucleinopathies. J Neurochem 2021;158(2):554– 568.

47. Benarroch EE, Schmeichel AM, Parisi JE. Depletion of mesopontine cholinergic and sparing of raphe neurons in multiple system atrophy. Neurology 2002;59(6):944–946.

48. Coon EA, Cutsforth-Gregory JK, Benarroch EE. Neuropathology of autonomic dysfunction in synucleinopathies. Movement Disorders 2018;33(3):349–358.

49. Gilman S, Koeppe RA, Nan B, et al. Cerebral cortical and subcortical cholinergic deficits in parkinsonian syndromes. Neurology 2010;74(18):1416–1423.

50. Chou KL, Dayalu P, Koeppe RA, et al. Serotonin Transporter Imaging in Multiple System Atrophy and Parkinson’s Disease. Mov Disord 2022;37(11):2301–2307.

