## Supplementary figures for "Atrophy in multiple system atrophy relates to mitochondrial and oligodendrocytic processes"

**FIGURE S1.** **Cortical surface vertex-wise analysis**

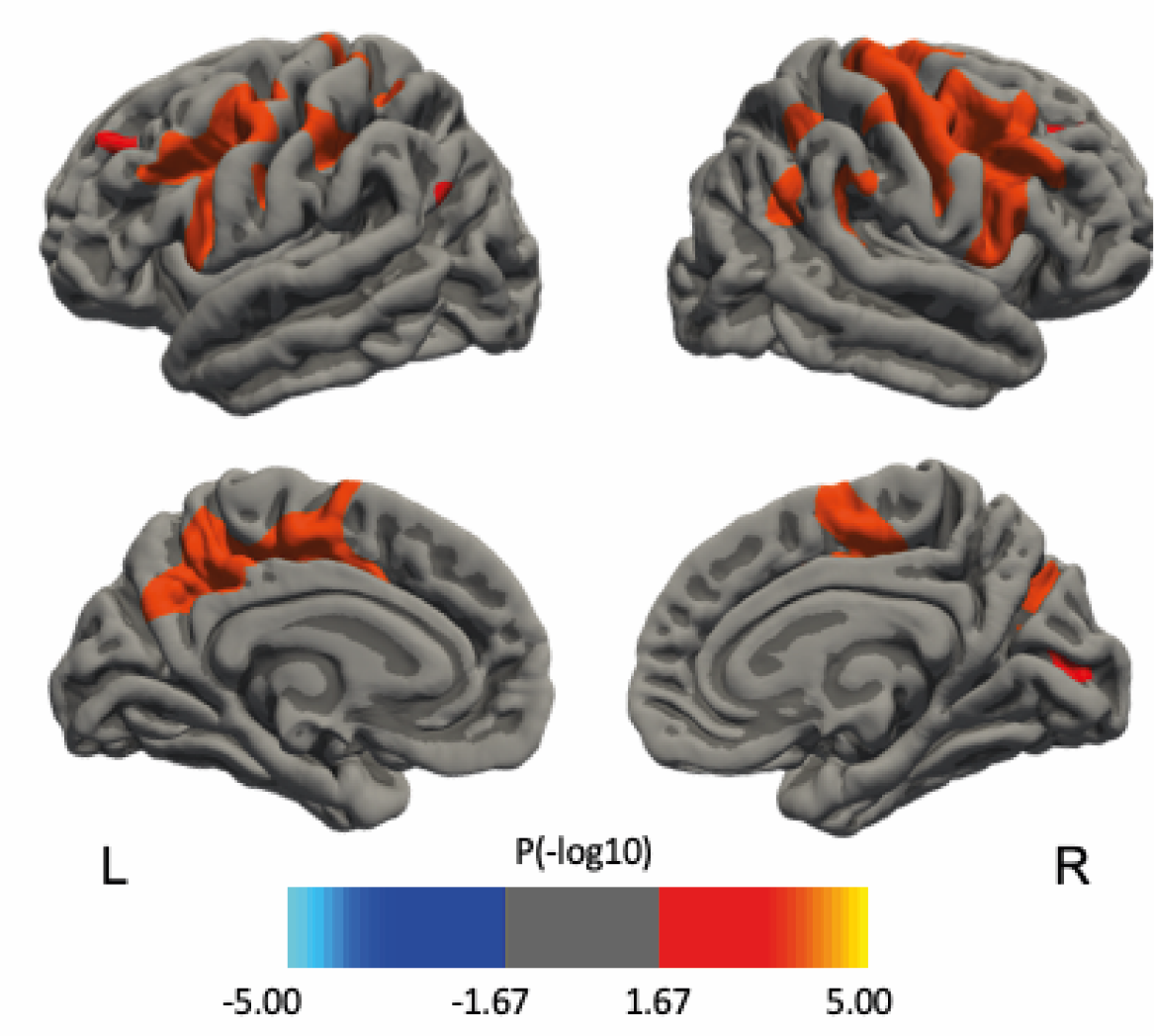

Lateral (upper row) and medial (lower row) sagittal views of the right and left hemispheres are shown. Clusters of reduced thickness in MSA patients compared to controls (in orange) were considered significant under a cluster-level P<0.05 using Monte-Carlo simulations and a voxel-level P value set at -log10 P value of 4.0 (P<0.0001) to minimize false positives.

*Abbreviations: L, left; R, right.*

**Figure S2: Specificity analysis: comparison with PD**

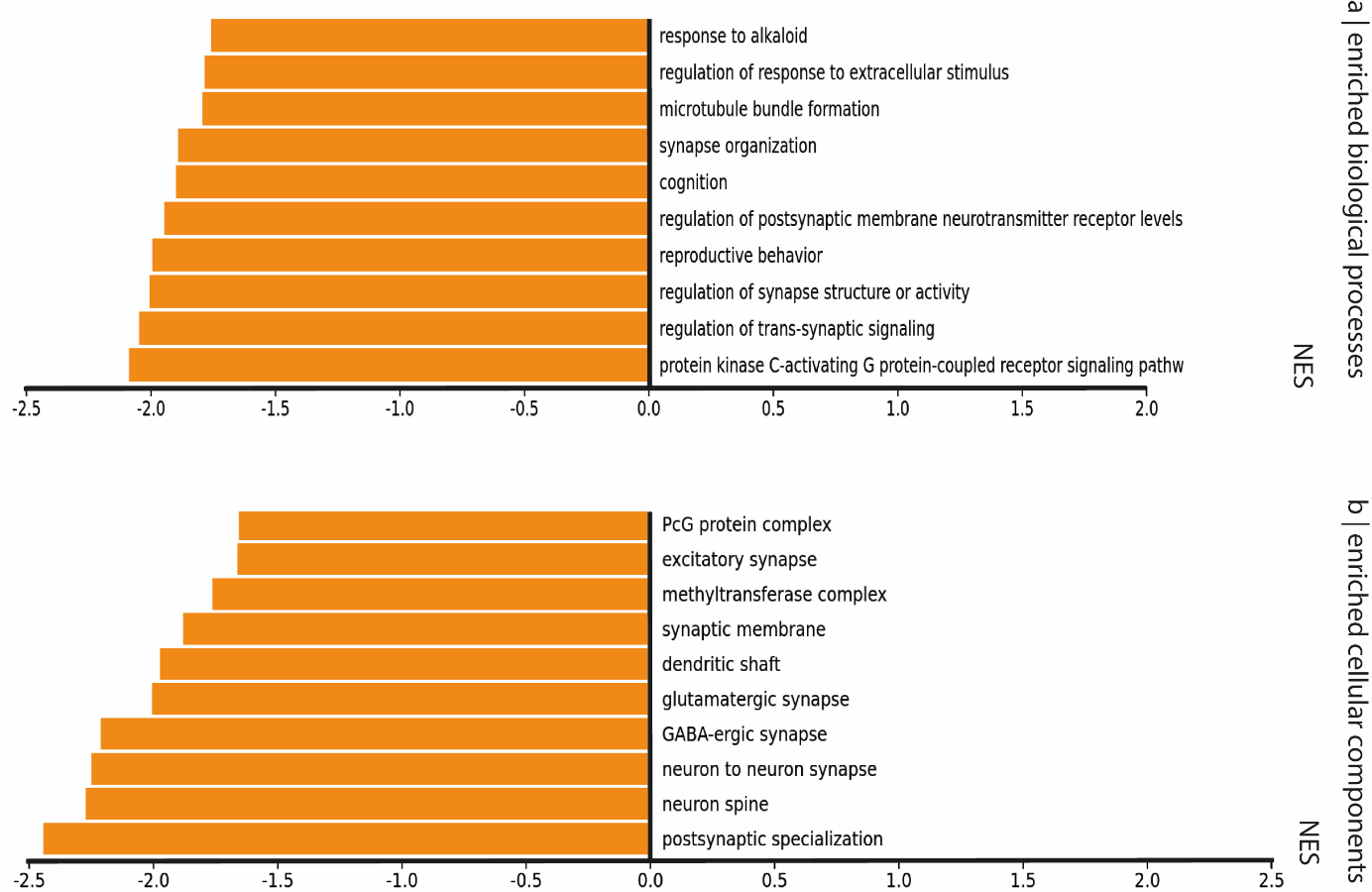

To test whether our gene enrichment findings in MSA were disease-specific, we replicated the same analysis on a sample of 57 patients with PD and 57 age- and sex-matched healthy controls recruited through the Quebec Parkinson Network (QPN), and scanned on a 3T Siemens PRISMA machine at the Montreal Neurological Institute-MNI, Montreal.(32) These participants were age- and sex-matched with MSA patients. Each participant’s T1-weighted image was segmented using the same image processing pipeline than described previously. A W-scoring approach was applied to the extracted values to remove the effects of age and sex using data from the control group. There was no significant deep brain atrophy in PD patients compared to controls (P>0.05).

PLS regression was performed between the two matrices: a matrix X of atrophy (57 patients by 50 regions) and a matrix Y of regional gene expression (15,611 genes by 50 regions). Of the 5 gene expression latent variables returned, two showed significance and explained 12.4% (LV4) and 7.6% (LV5) of the covariance in atrophy against random (LV4: P=0.06; LV5: P=0.02) and spatial null models (LV3: P=0.04; LV5: P=0.01). For both variables, regions with lower W-scores (more atrophy) had lower weights (LV4: r=0.35, P=0.01; LV5: r=0.28, P=0.05).

In terms of biological processes, regions associated with negatively-weighted genes in MSA were enriched for processes related to synaptic functions including protein kinase C-activating G protein-coupled receptor signaling pathway, regulation of transynaptic signaling, regulation of synapse structure or activity, regulation of postsynaptic membrane neurotransmitter receptor levels, and synapse organization. In terms of cellular components, we also found terms related to synapses, such as postsynaptic specialization, neuron spine, neuron-to-neuron synapse, GABA-ergic synapse. There was no significant enrichment of cell types, including oligodendrocytes.

*Abbreviations: NES, normalized enrichment score.*
