## Supplementary tables for "Atrophy in multiple system atrophy relates to mitochondrial and oligodendrocytic processes"

**TABLE S1. MRI acquisition parameters**

|  | **3T TRIO** | **1.5 OPTIMA** | **3T SIGNA** | **3T SKYRA** |
| --- | --- | --- | --- | --- |
| Magnetic field (Tesla) | 3 | 1.5 | 3 | 3 |
| Vendor | Siemens | GE | GE | Siemens |
| Head coil | 32 | 8 | 8 | 64 |
| Sequence type | SPGR | SPGR | SPGR | MPRAGE |
| TE (ms) | 2.94 | 1 | 2.788 | 2.34 |
| TR (ms) | 2200 | 9.356 | 6.564 | 2100 |
| TI (ms) | 900 | 300 | 400 | 900 |
| Flip angle | 10° | 15° | 11° | 8° |
| Voxel size (mm) | 1x1x1 | 0.9x0.9x1 | 1x1x1 | 0.9x0.9x0.9 |
| Number of averages | 1 | 1 | 1 | 1 |
| Spacing between slices | 1 | 1 | 1 | 1 |

MSA patients from the Genepark and BBBIPPS research protocols were scanned on the 3T Siemens TRIO MRI machine. MSA patients from the clinic cohort were scanned on the 3T GE SIGNA, 3T Siemens SKYRA, and 1.5T GE OPTIMA MRI machines. See [www.ppmi-info.org](http://www.ppmi-info.org) for the PPMI imaging protocols.

*Abbreviations: GE, General Electrics; MPRAGE, Magnetization Prepared - RApid Gradient Echo; MSA, multiple system atrophy; PPMI, Parkinson’s Progression Markers Initiative; SPGR, Spoiled gradient recalled echo; T, Tesla.*

**TABLE S2. Distribution of participants across the different MRI scanners**

|  | **3T Siemens TRIO^1^** | **1.5 GE OPTIMA^2^** | **3T GE SIGNA^2^** | **3T Siemens SKYRA^2^** | **PPMI 1.5T^3^** | **PPMI 3T^3^** | **Total** |
| --- | --- | --- | --- | --- | --- | --- | --- |
| **MSA** | 20 | 5 | 17 | 23 | 0 | 0 | 65 |
| **Controls** | 60 | 0 | 19 | 22 | 22 | 58 | 181 |

^1^ For the participants from the Genepark and BBBIPPS cohorts recruited at the Paris Brain Institute (ICM)

^2^ For the participants from the clinical cohort recruited at the movement disorders clinic at the Pitié-Salpêtrière University Hospital

^3^ For the participants from the PPMI cohort

*Abbreviations: GE, General Electric; MSA, multiple system atrophy; PPMI, Parkinson’s Progression Markers Initiative.*

**TABLE S3. W-scores associated with cortical and deep brain regions in the MSA group**

| **Regions** | **Mean W scores** | **T value** | **P** | **P_FDR_** | **Significance** |
| --- | --- | --- | --- | --- | --- |
| Left cerebellar WM | -2.67 | -12.10 | <0.0001 | <0.0001 | *** |
| Right cerebellar WM | -2.65 | -12.16 | <0.0001 | <0.0001 | *** |
| Pons | -2.52 | -13.83 | <0.0001 | <0.0001 | *** |
| Right putamen | -2.39 | -13.51 | <0.0001 | <0.0001 | *** |
| Right superior olive | -2.37 | -11.71 | <0.0001 | <0.0001 | *** |
| Left superior olive | -2.34 | -11.76 | <0.0001 | <0.0001 | *** |
| Left putamen | -2.18 | -12.44 | <0.0001 | <0.0001 | *** |
| Left SN | -1.88 | -11.39 | <0.0001 | <0.0001 | *** |
| Left inferior olive | -1.84 | -10.65 | <0.0001 | <0.0001 | *** |
| Right SN | -1.79 | -10.50 | <0.0001 | <0.0001 | *** |
| Right inferior olive | -1.67 | -10.49 | <0.0001 | <0.0001 | *** |
| Right lobule VIIb | -1.65 | -11.49 | <0.0001 | <0.0001 | *** |
| Left lobule I-IV | -1.62 | -9.64 | <0.0001 | <0.0001 | *** |
| Midbrain | -1.60 | -11.20 | <0.0001 | <0.0001 | *** |
| Vermis VIII | -1.56 | -11.08 | <0.0001 | <0.0001 | *** |
| Left lobule VIIb | -1.55 | -10.57 | <0.0001 | <0.0001 | *** |
| Left lobule Crus I | -1.54 | -8.98 | <0.0001 | <0.0001 | *** |
| Right lobule I-IV | -1.54 | -9.79 | <0.0001 | <0.0001 | *** |
| Left lobule VIIIa | -1.42 | -8.48 | <0.0001 | <0.0001 | *** |
| SCP | -1.33 | -10.39 | <0.0001 | <0.0001 | *** |
| Right lobule VIIIa | -1.33 | -8.46 | <0.0001 | <0.0001 | *** |
| Vermis IX | -1.32 | -9.32 | <0.0001 | <0.0001 | *** |
| Right lobule Crus I | -1.30 | -7.95 | <0.0001 | <0.0001 | *** |
| Vermis VI | -1.27 | -9.38 | <0.0001 | <0.0001 | *** |
| Left caudate | -1.26 | -8.06 | <0.0001 | <0.0001 | *** |
| Left pallidum | -1.25 | -7.72 | <0.0001 | <0.0001 | *** |
| Right precentral | -1.16 | -7.29 | <0.0001 | <0.0001 | *** |
| Right pars opercularis | -1.16 | -7.06 | <0.0001 | <0.0001 | *** |
| Right lobule VIIIb | -1.15 | -7.01 | <0.0001 | <0.0001 | *** |
| Right lobule IX | -1.13 | -8.70 | <0.0001 | <0.0001 | *** |
| Right caudate | -1.11 | -8.62 | <0.0001 | <0.0001 | *** |
| Right lobule VI | -1.09 | -8.47 | <0.0001 | <0.0001 | *** |
| Right lobule Crus II | -1.08 | -8.19 | <0.0001 | <0.0001 | *** |
| Left lobule VIIIb | -1.03 | -6.36 | <0.0001 | <0.0001 | *** |
| Left lobule VI | -1.03 | -7.88 | <0.0001 | <0.0001 | *** |
| Left lobule IX | -1.01 | -8.18 | <0.0001 | <0.0001 | *** |
| Left lobule Crus II | -1.00 | -7.00 | <0.0001 | <0.0001 | *** |
| Right pallidum | -0.97 | -6.63 | <0.0001 | <0.0001 | *** |
| Left lobule X | -0.96 | -6.88 | <0.0001 | <0.0001 | *** |
| Medulla | -0.96 | -7.73 | <0.0001 | <0.0001 | *** |
| Left precuneus | -0.92 | -6.09 | <0.0001 | <0.0001 | *** |
| Right lobule X | -0.92 | -6.05 | <0.0001 | <0.0001 | *** |
| Left thalamus | -0.90 | -7.95 | <0.0001 | <0.0001 | *** |
| Left precentral | -0.88 | -5.66 | <0.0001 | <0.0001 | *** |
| Right thalamus | -0.85 | -7.02 | <0.0001 | <0.0001 | *** |
| Vermis X | -0.84 | -6.32 | <0.0001 | <0.0001 | *** |
| Right caudal middle frontal | -0.82 | -5.42 | <0.0001 | <0.0001 | *** |
| Left caudal middle frontal | -0.79 | -5.24 | <0.0001 | <0.0001 | *** |
| Left lobule V | -0.76 | -5.59 | <0.0001 | <0.0001 | *** |
| Left inferior parietal | -0.75 | -5.95 | <0.0001 | <0.0001 | *** |
| Right supramarginal | -0.75 | -5.79 | <0.0001 | <0.0001 | *** |
| Right lobule V | -0.75 | -5.01 | <0.0001 | <0.0001 | *** |
| Right accumbens | -0.75 | -7.69 | <0.0001 | <0.0001 | *** |
| Left paracentral | -0.72 | -5.02 | <0.0001 | <0.0001 | *** |
| Left rostral middle frontal | -0.71 | -5.75 | <0.0001 | <0.0001 | *** |
| Left pars opercularis | -0.69 | -4.75 | <0.0001 | <0.0001 | *** |
| Right inferior parietal | -0.68 | -4.79 | <0.0001 | <0.0001 | *** |
| Right precuneus | -0.68 | -5.10 | <0.0001 | <0.0001 | *** |
| Left postcentral | -0.67 | -5.91 | <0.0001 | <0.0001 | *** |
| Left supramarginal | -0.67 | -4.90 | <0.0001 | <0.0001 | *** |
| Right superior parietal | -0.64 | -5.06 | <0.0001 | <0.0001 | *** |
| Right paracentral | -0.64 | -4.81 | <0.0001 | <0.0001 | *** |
| Left accumbens | -0.64 | -5.30 | <0.0001 | <0.0001 | *** |
| Left superior parietal | -0.61 | -4.47 | <0.0001 | <0.0001 | *** |
| Left superior frontal | -0.59 | -3.93 | 0.0002 | 0.0003 | *** |
| Right superior frontal | -0.57 | -4.05 | 0.0001 | 0.0002 | *** |
| Right pericalcarine | -0.57 | -5.11 | <0.0001 | <0.0001 | *** |
| Right pars triangularis | -0.56 | -4.31 | 0.0001 | 0.0002 | *** |
| Left pars triangularis | -0.53 | -3.39 | 0.0012 | 0.0017 | ** |
| Right banks superior temporal | -0.53 | -4.13 | 0.0001 | 0.0002 | *** |
| Right lateral occipital | -0.52 | -4.56 | <0.0001 | <0.0001 | *** |
| Right cuneus | -0.52 | -4.42 | <0.0001 | <0.0001 | *** |
| Right transverse temporal | -0.51 | -3.51 | 0.001 | 0.001 | ** |
| Left isthmus cingulate | -0.50 | -3.82 | <0.0001 | 0.001 | *** |
| Right rostral middle frontal | -0.50 | -3.71 | <0.0001 | 0.001 | *** |
| Left cuneus | -0.49 | -3.72 | <0.0001 | 0.001 | *** |
| Right postcentral | -0.47 | -4.05 | <0.0001 | 0.000 | *** |
| Left lingual | -0.47 | -3.84 | <0.0001 | 0.001 | *** |
| Right insula | -0.46 | -3.54 | 0.001 | 0.001 | ** |
| Left lateral orbitofrontal | -0.45 | -2.88 | 0.005 | 0.007 | ** |
| Right hippocampus | -0.45 | -3.68 | 0.001 | 0.001 | *** |
| Left middle temporal | -0.44 | -3.98 | <0.0001 | <0.0001 | *** |
| Left hippocampus | -0.42 | -4.04 | <0.0001 | <0.0001 | *** |
| Left banks superior temporal | -0.41 | -3.60 | 0.001 | 0.001 | *** |
| Right lateral orbitofrontal | -0.39 | -2.78 | 0.007 | 0.010 | ** |
| Right entorhinal | 0.37 | 3.01 | 0.004 | 0.005 | ** |
| Left pericalcarine | -0.37 | -3.36 | 0.001 | 0.002 | ** |
| Left posterior cingulate | -0.32 | -2.59 | 0.012 | 0.016 | * |
| Right pars orbitalis | -0.32 | -2.43 | 0.018 | 0.024 | * |
| Left superior temporal | -0.32 | -2.21 | 0.030 | 0.039 | * |
| Right superior temporal | -0.31 | -2.12 | 0.038 | 0.047 | * |
| Right frontal pole | -0.31 | -2.47 | 0.016 | 0.022 | * |
| Left frontal pole | -0.27 | -2.34 | 0.022 | 0.029 | * |
| Left amygdala | -0.27 | -2.33 | 0.023 | 0.030 | * |
| Right inferior temporal | -0.26 | -1.82 | 0.073 | 0.085 | ns |
| Right lingual | -0.26 | -2.20 | 0.031 | 0.039 | * |
| Right medial orbitofrontal | -0.26 | -1.91 | 0.060 | 0.071 | ns |
| Left lateral occipital | -0.25 | -2.05 | 0.045 | 0.055 | ns |
| Left transverse temporal | -0.24 | -1.92 | 0.060 | 0.071 | ns |
| Right amygdala | -0.24 | -1.96 | 0.055 | 0.066 | ns |
| Left insula | -0.23 | -1.59 | 0.117 | 0.136 | ns |
| Left entorhinal | 0.22 | 2.11 | 0.039 | 0.048 | * |
| Left medial orbitofrontal | -0.19 | -1.35 | 0.183 | 0.210 | ns |
| Right isthmus cingulate | -0.18 | -1.16 | 0.250 | 0.276 | ns |
| Right middle temporal | -0.18 | -1.20 | 0.236 | 0.262 | ns |
| Left pars orbitalis | -0.17 | -1.09 | 0.282 | 0.305 | ns |
| Left temporal pole | -0.16 | -1.29 | 0.201 | 0.228 | ns |
| Left inferior temporal | -0.15 | -1.21 | 0.232 | 0.261 | ns |
| Left caudal anterior cingulate | 0.14 | 1.11 | 0.270 | 0.295 | ns |
| Right parahippocampal | -0.11 | -0.74 | 0.460 | 0.493 | ns |
| Right fusiform | -0.08 | -0.58 | 0.563 | 0.598 | ns |
| Left rostral anterior cingulate | -0.07 | -0.52 | 0.607 | 0.640 | ns |
| Left parahippocampal | -0.06 | -0.39 | 0.696 | 0.715 | ns |
| Right temporal pole | -0.05 | -0.42 | 0.673 | 0.696 | ns |
| Right posterior cingulate | -0.05 | -0.43 | 0.669 | 0.696 | ns |
| Left fusiform | -0.03 | -0.24 | 0.815 | 0.829 | ns |
| Right caudal anterior cingulate | 0.01 | 0.06 | 0.955 | 0.963 | ns |
| Right rostral anterior cingulate | 0.00 | 0.01 | 0.991 | 0.991 | ns |

Mean regional W-scores across the entire MSA population were ranked from lowest (indicating greater atrophy) to highest (no atrophy) and compared to a null distribution using repeated one-sample t-tests, followed by an FDR correction for multiple comparisons. “lobule” refers to the cerebellum. ***P < 0.001; **0.001 < P ≤ 0.01; *0.001 < P ≤ 0.05

*Abbreviations: ns, non-significant; SCP, superior cerebellar peduncles; SN, substantia nigra; WM, white matter*

**TABLE S4. Cortical surface vertex-wise analysis**

| **Cluster** | **Max** | **VtxMax** | **Size(mm^2^)** | **MNI X** | **MNI Y** | **MNI Z** | **CWP** | **CWPLow** | **CWPHi** | **NVtxs** | **WghtVtx** | **Annotation** |
| --- | --- | --- | --- | --- | --- | --- | --- | --- | --- | --- | --- | --- |
| 1 | 9.6431 | 41178 | 4861.58 | -22.4 | -30.3 | 52.8 | 0.00020 | 0.00000 | 0.00040 | 10179 | 53926.14 | precentral |
| 2 | 8.1199 | 19602 | 3254.58 | -8.8 | -13.0 | 50.0 | 0.00020 | 0.00000 | 0.00040 | 7749 | 41835.11 | superior frontal |
| 3 | 6.9207 | 4490 | 1600.43 | -43.8 | -30.6 | 34.1 | 0.00020 | 0.00000 | 0.00040 | 3925 | 19592.87 | supramarginal |
| 4 | 4.7534 | 50524 | 215.61 | -24.2 | 40.4 | 32.3 | 0.00818 | 0.00659 | 0.00978 | 334 | 1407.30 | rostral middle frontal |
| 5 | 5.0366 | 131802 | 163.62 | -47.1 | -59.4 | 28.7 | 0.01832 | 0.01594 | 0.02069 | 342 | 1477.79 | inferior parietal |
| 6 | 4.4224 | 126197 | 161.96 | -32.9 | -63.4 | 39.7 | 0.01851 | 0.01613 | 0.02089 | 335 | 1351.49 | inferior parietal |

left hemisphere

right hemisphere

| **Cluster** | **Max** | **VtxMax** | **Size(mm^2^)** | **MNI X** | **MNI Y** | **MNI Z** | **CWP** | **CWPLow** | **CWPHi** | **NVtxs** | **WghtVtx** | **Annotation** |
| --- | --- | --- | --- | --- | --- | --- | --- | --- | --- | --- | --- | --- |
| 1 | 12.8666 | 8988 | 8634.24 | 33.5 | 8.6 | 33.0 | 0.00020 | 0.00000 | 0.00040 | 18075 | 106067.03 | caudal middle frontal |
| 2 | 10.2357 | 4675 | 1263.61 | 30.8 | -40.1 | 42.8 | 0.00020 | 0.00000 | 0.00040 | 3374 | 18684.06 | superior parietal |
| 3 | 5.3392 | 35270 | 649.56 | 19.5 | -74.3 | 34.7 | 0.00020 | 0.00000 | 0.00040 | 957 | 4098.40 | superior parietal |
| 4 | 6.4819 | 74861 | 620.10 | 49.4 | -36.2 | 16.1 | 0.00020 | 0.00000 | 0.00040 | 1473 | 7021.77 | superior temporal |
| 5 | 6.2097 | 33766 | 507.38 | 45.1 | -53.2 | 27.3 | 0.00020 | 0.00000 | 0.00040 | 1125 | 5386.30 | inferior parietal |
| 6 | 5.2718 | 156740 | 281.42 | 15.7 | -85.2 | 6.1 | 0.00160 | 0.00100 | 0.00240 | 447 | 1954.04 | pericalcarine |
| 7 | 4.9835 | 45260 | 181.58 | 21.7 | 36.0 | 32.4 | 0.01117 | 0.00938 | 0.01316 | 367 | 1568.43 | superior frontal |

Clusters of reduced thickness in MSA patients compared to controls (in orange) were considered significant under a cluster-level P<0.05 using Monte-Carlo simulations and a voxel-level P value set at -log_10_ P value of 4.0 (P<0.0001) to minimize false positives.

*Abbreviations: Max, Maximum value of t-statistic or z-score;CWP, Cluster-wise P value; CWPLow, Lower bound of cluster-wise P value; CWPHi, Upper bound of cluster-wise P value; MNIX, MNIY, MNIZ, peak coordinates (x, y, z) in the Montreal Neurological Institute space; NVtxs, Number of vertices in the cluster; Size (mm²), Size of the cluster in square millimeters; VtxMax, Vertex with maximum value; WghtVtx, Weighted vertex value (average statistic weighted by cluster size).*

**TABLE S5. W-scores associated with cortical and deep brain regions in the MSA subgroups**

| **Regions** | **Mean W score in MSAp** | **Mean W score in MSAc** | **Direction** | **T value** | **P** | **P_FDR_** | **Significance** |
| --- | --- | --- | --- | --- | --- | --- | --- |
| right putamen | -3.08 | -1.55 | MSAp < MSAc | -4.64 | <0.0001 | <0.0001 | *** |
| left putamen | -2.77 | -1.39 | MSAp < MSAc | -4.18 | <0.0001 | 0.001 | *** |
| left caudate | -1.80 | -0.76 | MSAp < MSAc | -3.57 | 0.001 | 0.006 | ** |
| pons | -1.69 | -3.33 | MSAp > MSAc | 5.17 | <0.0001 | <0.0001 | *** |
| left cerebellar WM | -1.64 | -3.87 | MSAp > MSAc | 5.97 | <0.0001 | <0.0001 | *** |
| right superior olive | -1.64 | -3.02 | MSAp > MSAc | 3.45 | 0.001 | 0.008 | ** |
| left superior olive | -1.61 | -2.99 | MSAp > MSAc | 3.51 | 0.001 | 0.007 | ** |
| left pallidum | -1.58 | -0.92 | MSAp < MSAc | -2.10 | 0.041 | 0.131 | ns |
| right caudate | -1.51 | -0.83 | MSAp < MSAc | -2.71 | 0.010 | 0.036 | * |
| left SN | -1.49 | -2.29 | MSAp > MSAc | 2.31 | 0.026 | 0.089 | ns |
| right precentral | -1.49 | -0.73 | MSAp < MSAc | -2.22 | 0.032 | 0.107 | ns |
| right cerebellar WM | -1.48 | -3.97 | MSAp > MSAc | 7.71 | <0.0001 | <0.0001 | *** |
| left inferior olive | -1.43 | -2.18 | MSAp > MSAc | 2.05 | 0.048 | 0.144 | ns |
| right SN | -1.38 | -2.27 | MSAp > MSAc | 2.33 | 0.026 | 0.089 | ns |
| right inferior olive | -1.28 | -1.97 | MSAp > MSAc | 2.05 | 0.048 | 0.144 | ns |
| right pallidum | -1.25 | -0.71 | MSAp < MSAc | -1.73 | 0.091 | 0.224 | ns |
| midbrain | -1.23 | -2.11 | MSAp > MSAc | 2.95 | 0.005 | 0.021 | * |
| right pars opercularis | -1.19 | -0.93 | MSAp < MSAc | -0.74 | 0.463 | 0.666 | ns |
| right lobule VIIb | -1.15 | -2.18 | MSAp > MSAc | 3.51 | 0.001 | 0.007 | ** |
| left lobule I-IV | -1.11 | -2.21 | MSAp > MSAc | 3.12 | 0.003 | 0.015 | * |
| left precentral | -1.06 | -0.71 | MSAp < MSAc | -1.01 | 0.317 | 0.534 | ns |
| left lobule VIIb | -1.04 | -2.10 | MSAp > MSAc | 3.30 | 0.002 | 0.011 | * |
| vermis VIII | -1.03 | -2.13 | MSAp > MSAc | 3.76 | 0.001 | 0.005 | ** |
| right caudal middle frontal | -1.02 | -0.59 | MSAp < MSAc | -1.28 | 0.206 | 0.426 | ns |
| SCP | -0.97 | -1.77 | MSAp > MSAc | 3.09 | 0.003 | 0.015 | * |
| Medulla | -0.96 | -1.06 | MSAp > MSAc | 0.37 | 0.715 | 0.861 | ns |
| left caudal middle frontal | -0.94 | -0.64 | MSAp < MSAc | -0.86 | 0.396 | 0.612 | ns |
| left pars opercularis | -0.93 | -0.57 | MSAp < MSAc | -1.07 | 0.293 | 0.508 | ns |
| right lobule I-IV | -0.92 | -2.26 | MSAp > MSAc | 4.65 | <0.0001 | <0.0001 | *** |
| left precuneus | -0.90 | -0.93 | MSAp > MSAc | 0.11 | 0.916 | 0.947 | ns |
| right lobule VIIIa | -0.89 | -1.87 | MSAp > MSAc | 2.87 | 0.006 | 0.024 | * |
| left paracentral | -0.85 | -0.47 | MSAp < MSAc | -1.14 | 0.260 | 0.479 | ns |
| left thalamus | -0.85 | -1.08 | MSAp > MSAc | 0.85 | 0.402 | 0.612 | ns |
| vermis IX | -0.85 | -1.98 | MSAp > MSAc | 4.24 | <0.0001 | 0.001 | *** |
| left lobule Crus I | -0.84 | -2.44 | MSAp > MSAc | 5.00 | <0.0001 | 0.000 | *** |
| right accumbens | -0.84 | -0.59 | MSAp < MSAc | -1.17 | 0.249 | 0.466 | ns |
| left inferior parietal | -0.84 | -0.87 | MSAp > MSAc | 0.12 | 0.904 | 0.944 | ns |
| right inferior parietal | -0.84 | -0.53 | MSAp < MSAc | -0.95 | 0.346 | 0.560 | ns |
| left lobule VIIIa | -0.84 | -2.20 | MSAp > MSAc | 3.72 | 0.001 | <0.0001 | ** |
| left rostral middle frontal | -0.82 | -0.58 | MSAp < MSAc | -0.87 | 0.392 | 0.612 | ns |
| vermis VI | -0.82 | -1.83 | MSAp > MSAc | 4.07 | <0.0001 | 0.002 | ** |
| right paracentral | -0.80 | -0.40 | MSAp < MSAc | -1.37 | 0.177 | 0.373 | ns |
| right lobule Crus II | -0.78 | -1.33 | MSAp > MSAc | 1.82 | 0.078 | 0.199 | ns |
| right supramarginal | -0.77 | -0.67 | MSAp < MSAc | -0.32 | 0.753 | 0.871 | ns |
| left supramarginal | -0.77 | -0.74 | MSAp < MSAc | -0.09 | 0.931 | 0.947 | ns |
| right lobule IX | -0.75 | -1.57 | MSAp > MSAc | 3.48 | 0.001 | 0.007 | ** |
| left postcentral | -0.75 | -0.59 | MSAp < MSAc | -0.60 | 0.553 | 0.733 | ns |
| left lobule Crus II | -0.75 | -1.16 | MSAp > MSAc | 1.22 | 0.229 | 0.445 | ns |
| left superior parietal | -0.73 | -0.55 | MSAp < MSAc | -0.59 | 0.560 | 0.734 | ns |
| left superior frontal | -0.73 | -0.40 | MSAp < MSAc | -0.96 | 0.341 | 0.560 | ns |
| right precuneus | -0.72 | -0.62 | MSAp < MSAc | -0.35 | 0.731 | 0.871 | ns |
| right superior frontal | -0.71 | -0.34 | MSAp < MSAc | -1.17 | 0.247 | 0.466 | ns |
| right thalamus | -0.69 | -1.22 | MSAp > MSAc | 1.93 | 0.061 | 0.168 | ns |
| left pars triangularis | -0.68 | -0.35 | MSAp < MSAc | -0.95 | 0.345 | 0.560 | ns |
| right superior parietal | -0.68 | -0.58 | MSAp < MSAc | -0.34 | 0.738 | 0.871 | ns |
| left accumbens | -0.68 | -0.56 | MSAp < MSAc | -0.43 | 0.672 | 0.826 | ns |
| right lateral occipital | -0.67 | -0.44 | MSAp < MSAc | -1.05 | 0.299 | 0.511 | ns |
| right pars triangularis | -0.64 | -0.45 | MSAp < MSAc | -0.65 | 0.519 | 0.707 | ns |
| left lobule IX | -0.62 | -1.48 | MSAp > MSAc | 3.46 | 0.001 | 0.007 | ** |
| left isthmus cingulate | -0.62 | -0.15 | MSAp < MSAc | -1.57 | 0.125 | 0.293 | ns |
| left lobule VI | -0.62 | -1.58 | MSAp > MSAc | 3.91 | <0.0001 | 0.003 | ** |
| right lobule Crus I | -0.60 | -2.17 | MSAp > MSAc | 5.23 | <0.0001 | <0.0001 | *** |
| right lobule VI | -0.60 | -1.66 | MSAp > MSAc | 4.42 | <0.0001 | 0.001 | *** |
| right banks superior temporal | -0.60 | -0.46 | MSAp < MSAc | -0.47 | 0.644 | 0.811 | ns |
| left posterior cingulate | -0.55 | -0.01 | MSAp < MSAc | -1.99 | 0.054 | 0.158 | ns |
| vermis X | -0.54 | -1.30 | MSAp > MSAc | 2.98 | 0.005 | 0.019 | * |
| right transverse temporal | -0.54 | -0.43 | MSAp < MSAc | -0.30 | 0.769 | 0.881 | ns |
| right rostral middle frontal | -0.53 | -0.50 | MSAp < MSAc | -0.13 | 0.896 | 0.944 | ns |
| left lobule VIIIb | -0.53 | -1.90 | MSAp > MSAc | 4.39 | <0.0001 | 0.001 | *** |
| left lobule X | -0.52 | -1.49 | MSAp > MSAc | 3.21 | 0.003 | 0.012 | * |
| right pericalcarine | -0.50 | -0.66 | MSAp > MSAc | 0.66 | 0.513 | 0.707 | ns |
| right cuneus | -0.50 | -0.53 | MSAp > MSAc | 0.12 | 0.903 | 0.944 | ns |
| left transverse temporal | -0.49 | 0.10 | MSAp < MSAc | -2.18 | 0.035 | 0.113 | ns |
| right lobule V | -0.47 | -1.12 | MSAp > MSAc | 1.93 | 0.061 | 0.168 | ns |
| right postcentral | -0.46 | -0.38 | MSAp < MSAc | -0.32 | 0.749 | 0.871 | ns |
| right lobule X | -0.46 | -1.49 | MSAp > MSAc | 3.19 | 0.003 | 0.012 | * |
| right lobule VIIIb | -0.46 | -2.07 | MSAp > MSAc | 5.23 | <0.0001 | <0.0001 | *** |
| right insula | -0.44 | -0.42 | MSAp < MSAc | -0.07 | 0.942 | 0.950 | ns |
| right frontal pole | -0.43 | 0.07 | MSAp < MSAc | -1.97 | 0.056 | 0.161 | ns |
| left middle temporal | -0.42 | -0.70 | MSAp > MSAc | 1.10 | 0.280 | 0.493 | ns |
| right isthmus cingulate | -0.41 | 0.28 | MSAp < MSAc | -1.89 | 0.066 | 0.178 | ns |
| left lingual | -0.41 | -0.45 | MSAp > MSAc | 0.17 | 0.869 | 0.944 | ns |
| right parahippocampal | -0.40 | -0.16 | MSAp < MSAc | -0.78 | 0.443 | 0.645 | ns |
| left superior temporal | -0.40 | -0.48 | MSAp > MSAc | 0.23 | 0.817 | 0.918 | ns |
| left lobule V | -0.38 | -1.26 | MSAp > MSAc | 2.96 | 0.005 | 0.021 | * |
| left frontal pole | -0.38 | -0.06 | MSAp < MSAc | -1.43 | 0.158 | 0.353 | ns |
| left banks superior temporal | -0.36 | -0.34 | MSAp < MSAc | -0.09 | 0.927 | 0.947 | ns |
| left pericalcarine | -0.36 | -0.33 | MSAp < MSAc | -0.15 | 0.885 | 0.944 | ns |
| left hippocampus | -0.36 | -0.69 | MSAp > MSAc | 1.55 | 0.127 | 0.293 | ns |
| left fusiform | -0.35 | 0.05 | MSAp < MSAc | -1.26 | 0.217 | 0.435 | ns |
| right fusiform | -0.35 | 0.10 | MSAp < MSAc | -1.42 | 0.163 | 0.354 | ns |
| right pars orbitalis | -0.34 | -0.30 | MSAp < MSAc | -0.14 | 0.891 | 0.944 | ns |
| right hippocampus | -0.34 | -0.74 | MSAp > MSAc | 1.46 | 0.152 | 0.345 | ns |
| right lingual | -0.30 | -0.18 | MSAp < MSAc | -0.46 | 0.646 | 0.811 | ns |
| left lateral occipital | -0.29 | -0.43 | MSAp > MSAc | 0.54 | 0.591 | 0.757 | ns |
| right superior temporal | -0.29 | -0.56 | MSAp > MSAc | 0.83 | 0.414 | 0.618 | ns |
| left cuneus | -0.28 | -0.62 | MSAp > MSAc | 1.22 | 0.230 | 0.445 | ns |
| right inferior temporal | -0.26 | -0.51 | MSAp > MSAc | 0.78 | 0.439 | 0.645 | ns |
| left medial orbitofrontal | -0.24 | -0.05 | MSAp < MSAc | -0.57 | 0.571 | 0.740 | ns |
| left parahippocampal | -0.23 | -0.14 | MSAp < MSAc | -0.28 | 0.782 | 0.887 | ns |
| left lateral orbitofrontal | -0.22 | -0.69 | MSAp > MSAc | 1.27 | 0.213 | 0.432 | ns |
| right medial orbitofrontal | -0.20 | -0.20 | MSAp < MSAc | -0.01 | 0.994 | 0.994 | ns |
| right amygdala | -0.18 | -0.34 | MSAp > MSAc | 0.64 | 0.525 | 0.707 | ns |
| right lateral orbitofrontal | -0.16 | -0.67 | MSAp > MSAc | 1.56 | 0.126 | 0.293 | ns |
| left insula | -0.15 | -0.36 | MSAp > MSAc | 0.64 | 0.527 | 0.707 | ns |
| left pars orbitalis | -0.15 | -0.20 | MSAp > MSAc | 0.15 | 0.883 | 0.944 | ns |
| right posterior cingulate | -0.12 | 0.11 | MSAp < MSAc | -0.84 | 0.405 | 0.612 | ns |
| left inferior temporal | -0.12 | -0.50 | MSAp > MSAc | 1.41 | 0.165 | 0.354 | ns |
| right middle temporal | -0.07 | -0.44 | MSAp > MSAc | 1.10 | 0.278 | 0.493 | ns |
| left temporal pole | -0.07 | -0.52 | MSAp > MSAc | 1.75 | 0.087 | 0.217 | ns |
| right temporal pole | -0.04 | -0.31 | MSAp > MSAc | 0.93 | 0.358 | 0.571 | ns |
| right caudal anterior cingulate | -0.01 | 0.23 | MSAp < MSAc | -0.73 | 0.472 | 0.669 | ns |
| left amygdala | 0.00 | -0.50 | MSAp > MSAc | 1.84 | 0.076 | 0.199 | ns |
| right rostral anterior cingulate | 0.00 | 0.19 | MSAp < MSAc | -0.72 | 0.476 | 0.669 | ns |
| left rostral anterior cingulate | 0.02 | -0.10 | MSAp > MSAc | 0.38 | 0.705 | 0.857 | ns |
| left caudal anterior cingulate | 0.13 | 0.17 | MSAp < MSAc | -0.15 | 0.882 | 0.944 | ns |
| left entorhinal | 0.31 | 0.03 | MSAp > MSAc | 1.13 | 0.268 | 0.486 | ns |
| right entorhinal | 0.40 | 0.27 | MSAp > MSAc | 0.44 | 0.663 | 0.824 | ns |

Regional W-scores in the MSAp and MSAc subgroups were compared using two-sample t-tests followed by an FDR correction for multiple comparisons. “lobule” refers to the cerebellum. ***P < 0.001; **0.001 < P ≤ 0.01; *0.001 < P ≤ 0.05

*Abbreviations: MSA, multiple system atrophy; MSAp, parkinsonian variant of MSA; MSAc; cerebellar variant of MSA; ns, non-significant; SCP, superior cerebellar peduncles; SN, substantia nigra; WM, white matter*

**TABLE S6. Gene set enrichment analysis in the whole MSA sample** **using PANTHER**

| **Gene set** | **Description** | **Size** | **P** | **P_FDR_** |
| --- | --- | --- | --- | --- |
| GO:0006810 | Transport | 3015 | <0.0001 | <0.0001 |
| GO:0045333 | Cellular respiration | 171 | <0.0001 | <0.0001 |
| GO:0007005 | Mitochondrion organization | 408 | <0.0001 | <0.0001 |
| GO:0006119 | Oxidative phosphorylation | 100 | <0.0001 | <0.0001 |
| GO:0006091 | Generation of precursor metabolites and energy | 327 | <0.0001 | <0.0001 |
| GO:0044281 | Small molecule metabolic process | 1312 | <0.0001 | <0.0001 |
| GO:0051234 | Establishment of localization | 3231 | <0.0001 | <0.0001 |
| GO:0015980 | Energy derivation by oxidation of organic compounds | 237 | <0.0001 | <0.0001 |
| GO:0055085 | Transmembrane transport | 980 | <0.0001 | <0.0001 |
| GO:0009060 | Aerobic respiration | 142 | <0.0001 | <0.0001 |
| GO:0019646 | Aerobic electron transport chain | 72 | <0.0001 | <0.0001 |
| GO:0009150 | Purine ribonucleotide metabolic process | 317 | <0.0001 | <0.0001 |
| GO:0051179 | Localization | 3718 | <0.0001 | <0.0001 |
| GO:0022904 | Respiratory electron transport chain | 101 | <0.0001 | <0.0001 |
| GO:0009152 | Purine ribonucleotide biosynthetic process | 164 | <0.0001 | <0.0001 |

Significant biological processes associated the third latent variable obtained using PANTHER (version 19.0, 20240619).[1] Only results associated with negatively-weighted genes (i.e., genes more expressed in regions with greater atrophy) are reported.

*Abbreviations: FDR, false discovery rate; GO, gene-ontology.*

**Table S7: Gene set enrichment sub-analyses on MSA subgroups:**

**a | MSAp subgroup**

| **Gene Set** | **Biological processes** | **Size** | **Leading Edge Number** | **Enrichment score** | **Normalized enrichment score** | **P** | **P_FDR_** |
| --- | --- | --- | --- | --- | --- | --- | --- |
| GO:0010257 | NADH dehydrogenase complex assembly | 54 | 33 | -0.57 | -2.31 | <0.0001 | <0.0001 |
| GO:0006882 | intracellular zinc ion homeostasis | 28 | 16 | -0.64 | -2.27 | <0.0001 | <0.0001 |
| GO:0007229 | integrin-mediated signaling pathway | 91 | 55 | -0.48 | -2.18 | <0.0001 | <0.0001 |
| GO:1901342 | regulation of vasculature development | 222 | 109 | -0.40 | -2.09 | <0.0001 | 0.0031 |
| GO:0002274 | myeloid leukocyte activation | 165 | 87 | -0.42 | -2.07 | <0.0001 | 0.0036 |
| GO:0022900 | electron transport chain | 146 | 96 | -0.42 | -2.05 | <0.0001 | 0.0039 |
| GO:0009636 | response to toxic substance | 184 | 98 | -0.40 | -2.02 | <0.0001 | 0.0061 |
| GO:0015980 | energy derivation by oxidation of organic compounds | 280 | 144 | -0.38 | -2.01 | <0.0001 | 0.0058 |
| GO:0016053 | organic acid biosynthetic process | 257 | 106 | -0.38 | -1.97 | <0.0001 | 0.0071 |
| GO:0042060 | wound healing | 310 | 137 | -0.37 | -1.96 | <0.0001 | 0.0069 |

**b | MSAc subgroup**

| **Gene Set** | **Biological processes** | **Size** | **Leading Edge Number** | **Enrichment score** | **Normalized enrichment score** | **P** | **P_FDR_** |
| --- | --- | --- | --- | --- | --- | --- | --- |
| GO:0010257 | NADH dehydrogenase complex assembly | 54 | 35 | -0.65 | -2.51 | <0.0001 | <0.0001 |
| GO:1902600 | proton transmembrane transport | 114 | 59 | -0.56 | -2.47 | <0.0001 | <0.0001 |
| GO:0022900 | electron transport chain | 146 | 70 | -0.50 | -2.29 | <0.0001 | 0.0003 |
| GO:0007005 | mitochondrion organization | 439 | 207 | -0.43 | -2.21 | <0.0001 | 0.0003 |
| GO:0006839 | mitochondrial transport | 161 | 69 | -0.46 | -2.14 | <0.0001 | 0.0006 |
| GO:0072522 | purine-containing compound biosynthetic process | 216 | 87 | -0.44 | -2.11 | <0.0001 | 0.0007 |
| GO:0009141 | nucleoside triphosphate metabolic process | 223 | 114 | -0.43 | -2.08 | <0.0001 | 0.0007 |
| GO:0015980 | energy derivation by oxidation of organic compounds | 280 | 124 | -0.42 | -2.07 | <0.0001 | 0.0007 |
| GO:1901293 | nucleoside phosphate biosynthetic process | 240 | 92 | -0.42 | -2.02 | <0.0001 | 0.0013 |
| GO:0019693 | ribose phosphate metabolic process | 396 | 168 | -0.39 | -1.96 | <0.0001 | 0.0030 |

We ran secondary PLS regression using deep brain regions and GSEA analyses on MSAp and MSAc subgroups to identify which terms were specifically overexpressed in each subgroup.

For MSAp patients compared to controls, two latent variables of gene expression explained atrophy, each explaining respectively 32.2% and 8.8% of the covariance (against random nulls: both P=0.01; against spatial nulls: P=0.002 and P=0.003). In both latent variables, the regional weights were positively correlated with atrophy W-scores (r=0.44, P=0.001; r=0.30, P=0.04). In MSAc patients compared to controls, one latent variable explained 52.7% of the covariance in atrophy (against random nulls: P<0.0001; against spatial nulls: P=0.005). The regional weights of deep brain regions on this latent variable were positively correlated with W-scores of atrophy (r=0.73, P<0.0001). Similar to the results on the whole MSA sample, GSEA on negatively-weighted genes also revealed terms related to mitochondrial functions in both subgroups. For MSAp, there were additional terms such as intracellular zinc ion homeostasis (normalized enrichment score: -2.27, P_FDR_<0.0001) and regulation of vasculature development (-2.09, P_FDR_ =0.003).

**TABLE S8. Neural cell type gene enrichment analysis**

|  | **r** | **P** | **P_spatial_** | **P_FDR spatial_** |
| --- | --- | --- | --- | --- |
| **Oligodendrocytes** | -0.386 | 0.0057* | **0.002** | **0.003** |
| **OPC** | -0.114 | 0.432 | ~~--~~ | ~~--~~ |
| **Microglial cells** | -0.123 | 0.397 | ~~--~~ | ~~--~~ |
| **Astrocytes** | -0.047 | 0.748 | ~~--~~ | ~~--~~ |
| **Excitatory neurons** | -0.306 | 0.031 | 0.159 | ~~--~~ |
| **Inhibitory neurons** | -0.262 | 0.066 | ~~--~~ | ~~--~~ |
| **Endothelial cells** | -0.317 | 0.025 | **0.010** | **0.010** |

Pearson correlations between regional W-scores and gene expression across neural cell types in deep brain regions. A correlation was considered significant when its P value was below the Bonferroni-corrected threshold of P<0.007 (indicated by *) and when the FDR-corrected P value associated with the spatial null models was below 0.05 (in bold).

*Abbreviations: FDR, false discovery rate; OPC, oligodendrocyte progenitor cells; P_spatial_, P value associated with spatial null models; P_FDR_, FDR-corrected P value.*

| **Neurotransmitter** | | **r** | **P** | **P_spatial_** | **P_FDR spatial_** | **Significance** |
| --- | --- | --- | --- | --- | --- | --- |
| **Serotonin** | **5-HT_1A_** | 0.75 | <0.0001* | **<0.0001** | **<0.0001** | *** |
|  | **5-HT_1B_** | -0.10 | 0.332 | -- | -- | -- |
|  | **5-HT_2A_** | 0.36 | 0.001* | **0.023** | **0.046** | * |
|  | **5-HT_4_** | 0.15 | 0.155 | -- | -- | -- |
|  | **5-HT_6_** | 0.02 | 0.880 | -- | -- | -- |
|  | **5-HTT** | -0.14 | 0.184 | -- | -- | -- |
| **Acetylcholine** | **α4β2** | -0.61 | <0.0001* | **<0.0001** | **<0.0001** | *** |
|  | **VAChT** | -0.29 | 0.008 | -- | -- | ns |
|  | **M1** | 0.12 | 0.267 | -- | -- | -- |
| **Dopamine** | **D1** | 0.24 | 0.023 | -- | -- | ns |
|  | **D2** | 0.04 | 0.731 | -- | -- | -- |
|  | **DAT** | 0.01 | 0.954 | -- | -- | -- |
| **GABA** | **GABA_A_/BZ** | 0.42 | <0.0001* | **0.017** | **0.046** | * |
| **Histamine** | **H3** | -0.07 | 0.481 | -- | -- | -- |
| **Cannabinoids** | **CB1** | 0.24 | 0.023 | -- | -- | ns |
| **Opioïd** | **μ** | 0.14 | 0.183 | -- | -- | -- |
| **Noradrenaline** | **NET** | -0.36 | 0.001* | **0.026** | **0.046** | * |
| **Glutamate** | **NMDA** | 0.03 | 0.756 | -- | -- | -- |
|  | **mGluR5** | 0.28 | 0.007 | -- | -- | -- |

**TABLE S9. Neurotransmitter mapping**

Spearman correlations between regional W-scores and density values of receptors in cortical and deep brain regions. A correlation was considered significant when its P value was below the Bonferroni-corrected threshold of P<0.003 (indicated by *) and when the FDR-corrected P value associated with the spatial null models was below 0.05 (in bold).

*Abbreviations: FDR, false discovery rate; P_spatial_, P value associated with spatial null models; P_FDR_, FDR-corrected P value.*
